# Altered Cerebral Blood Flow in Older Adults with Alzheimer’s Disease: A Systematic Review

**DOI:** 10.1101/2022.03.24.22272916

**Authors:** Cecily G. Swinford, Shannon L. Risacher, Yu-Chien Wu, Liana G. Apostolova, Sujuan Gao, Andrew J. Saykin

## Abstract

The prevalence of Alzheimer’s disease is projected to reach 13 million in the U.S. by 2050. Although major efforts have been made to avoid this outcome, so far there are no treatments that can stop or reverse the progressive cognitive decline that defines Alzheimer’s disease. The utilization of preventative treatment before significant cognitive decline has occurred may ultimately be the solution, necessitating a reliable biomarker of preclinical/prodromal disease stages to determine which older adults are most at risk. Quantitative cerebral blood flow is a promising potential early biomarker for Alzheimer’s disease, but the spatiotemporal patterns of altered cerebral blood flow in Alzheimer’s disease are not fully understood. The current systematic review compiles the findings of 29 original studies that compared quantitative cerebral blood flow in older adults with mild cognitive impairment or Alzheimer’s disease to that of cognitively normal older adults and/or assessed the relationship between cerebral blood flow and objective cognitive function. Individuals with Alzheimer’s disease had relatively decreased cerebral blood flow in all brain regions investigated, especially the temporoparietal and posterior cingulate, while individuals with mild cognitive impairment had less consistent results, with relatively increased cerebral blood flow reported in the temporal lobe and thalamus. Most papers reported a positive correlation between cerebral blood flow and cognitive function. This review highlights the need for more studies comparing cerebral blood flow between cognitively normal individuals and those with mild cognitive impairment, as well as the importance of including potential confounding factors in these analyses.

## INTRODUCTION

Alzheimer’s disease (AD) is the leading cause of dementia; 60-80% of dementia cases are attributable to AD (Alzheimer’s Disease Facts and Figures, 2021). Both the prevalence and mortality rate of AD are increasing in the U.S. and globally, and there are no current treatments that can stop or reverse the progressive loss of cognitive function caused by AD (Alzheimer’s Disease Facts and Figures, 2021). Clinical trials of treatments targeting pathologic forms of amyloid-β and tau proteins in the brain have successfully reduced them but have been unable to stop the progression of cognitive decline (Zhu et al., 2020). In order to effectively treat AD, a preventative treatment administered before irreversible neuronal damage has occurred may be necessary (Zhu et al., 2020). An effective early biomarker for the identification of older adults who are most likely to develop AD is critical (Blennow & Zetterberg, 2018; Counts et al., 2017). The disease process of AD can begin decades before cognitive decline is apparent and can manifest as subjective cognitive decline (SCD) or mild cognitive impairment (MCI; Petersen et al., 1999; Petersen, 2016; Jessen et al., 2014; Jessen et al., 2020). MCI is a stage of decreased cognitive functioning measured on objective cognitive tests accompanied by the preserved ability to complete day-to-day activities (Petersen et al., 1999). SCD, during which individuals still perform in the normal range on objective cognitive tests but report a subjective decline from their normal cognitive state, has been described as an even earlier stage of AD (Jessen et al., 2014). An effective early biomarker for AD that can lead to preventative treatment must be able to distinguish individuals with SCD and MCI from normally aging individuals and from older adults experiencing cognitive changes due to other reasons, such as depression (Culpepper et. al., 2017).

Cerebral blood flow (CBF) is a potential early biomarker for AD. CBF is globally decreased in patients with AD compared to age-matched non-demented counterparts. It has been shown that CBF is altered in individuals with preclinical AD as well, and that patterns of altered CBF are correlated with disease severity and progression from one diagnostic stage to the next (Duan et al., 2021). Due to the brain’s sensitivity to changes in blood pressure, chronic hypertension leads to constriction of the cerebral vasculature. Additionally, blood vessels in the brain become more rigid with age, so the combination of chronic hypertension and aging lead to a hyper-constricted cerebral vasculature and ultimately chronically reduced CBF (Iadecola & Gottesman, 2019). Although the longstanding amyloid cascade hypothesis of AD posits that amyloid-β and its aggregation ultimately lead to the disease processes and outcomes in AD (Hardy & Higgins, 1992), the more recently suggested two-hit hypothesis states that cerebrovascular dysfunction and amyloid-β pathology culminate to initiate and propagate AD (Zlokovic, 2011).

CBF has been studied in the context of AD and other dementias for several decades and has been measured by a variety of neuroimaging methods over time. The first measurements of CBF in humans were made in 1948 using the inhalation of nitrous oxide and quantification of the inert gas in venous blood (Kety & Schmidt, 1948). In the 1960s, the use of radioactive tracers including krypton-85 and xenon-133 allowed for the measurement of regional as well as global CBF (Lassen & Ingvar, 1961; Lassen et al., 1963). The development of positron emission tomography (PET) soon led to the use of positron-emitting tracers, particularly oxygen-15, for CBF measurement (Jones et al., 1976). Dynamic contrast enhanced magnetic resonance imaging (DCE MRI) and dynamic susceptibility-contrast (DSC) MRI were developed around the same time as PET and utilized gadolinium-based contrast agents that had magnetic properties to trace and quantify CBF without the need for radioactive tracers. (Edelman et al., 1990; Rempp et al., 1994; Maeda et al., 1993).

The most common methods of the studies included in this review are single-photon emission computerized tomography (SPECT) and the more recently developed arterial spin labeled (ASL) MRI. SPECT requires the blood to be labeled with an injection of a radioactive tracer, such as iodine-123 or technetium-99m (Warwick, 2004). ASL MRI, on the other hand, is completely noninvasive. In ASL MRI, blood is labeled as it flows into the brain by inverting the magnetization of the water molecules in the blood and then comparing the labeled image to an unlabeled background image (Detre et al., 2012). ASL MRI can therefore be safely and comfortably used at multiple time points to monitor changes in CBF over the course of disease or treatment.

In order to utilize CBF as an early biomarker for AD, we need to better understand how CBF changes throughout the progression of AD, including in prodromal stages. The roles of other AD-related pathologies (amyloid-β, tau, gray matter atrophy, white matter hyperintensities), AD risk factors (apolipoprotein E (*APOE*) ε4 genotype, diabetes, hypertension, and other cardiovascular diseases), and demographic factors (age, sex, race/ethnicity, education, environmental risk factors) must also be taken into consideration when measuring CBF.

The purpose of this review is to consolidate the literature comparing CBF in older adults with MCI or AD to those that are cognitively normal (CN). Patterns of altered CBF will be summarized using results from several brain regions. Results will also be summarized for the relationship between cognitive exam scores and CBF in older adults. The aim is to compile and synthesize the results of the existing literature concerning altered CBF in AD. This is important because various methods of CBF measurement have been used over the years; standardization of results will help to clarify whether previous findings are consistent. A recently published review (Zhang et al., 2021) covers the same topic with a broader scope. It includes studies that measured CBF velocity, and it does not exclude articles based on the age of participants with AD. However, Zhang et al. (2021) do not include papers that report voxel-wise results in the form of z or t scores. The current review employs an age cutoff for the AD participants included because early onset AD, diagnosed before age 65, may be a different variant of AD; early onset AD patients tend to decline more rapidly and are more likely to have cognitive declines in areas other than memory as the initial symptoms. Therefore, we excluded papers where the mean age of AD participants minus two standard deviations (SDs) is less than 60, meaning that nearly 98% of the individuals studied in each paper are at least 60 years old. This defines the overall AD population as mostly late onset. We also included papers that report voxel-wise results to assess their consistency with papers that report regional CBF values.

The objective of this review is to assess the cumulative evidence of altered CBF in older adults with MCI and AD in multiple brain regions and of the relationship between CBF and cognitive exam scores in these individuals. We also aim to assess the consistency of findings across papers that use a variety of imaging, processing, and analytic methods. Our main question is one of correlation rather than causation between diagnosis and CBF. In reality, it is likely that AD pathologies and altered CBF exacerbate one another and have a cyclical relationship, and it is not clear which of the two is present first or whether both arise simultaneously from a common cause.

## METHODS

This systematic review includes all studies in which resting CBF is compared between diagnostic groups (CN, MCI, AD) and/or correlated with cognitive exam scores. Only studies in which the mean age of AD patients minus two SDs is at least 60 years old are included. If there was not a SD listed for age of participants with AD, that paper was not eligible unless the minimum age was at least 60. We chose to use a cutoff for participant age so that a majority of the AD patients would be over the age of 60 and less likely to have early-onset variants of AD. Both observational studies and clinical trials that measured CBF at rest prior to treatment are included, as well as both cross-sectional and longitudinal designs. Only manuscripts written in English were included. All manuscripts are published as original research papers or brief reports; conference abstracts were not included. There were no restrictions on the year of publication, and papers that used any established method for the quantification of CBF were eligible for inclusion. Studies that were ineligible either did not measure CBF at rest or did not use resting CBF as an outcome measure in multiple diagnostic groups and/or in comparison with cognitive test scores.

PubMed was searched on March 23, 2021, through the National Library of Medicine and with access via Indiana University School of Medicine. 1197 potential articles resulted from a PubMed search of “brain-blood supply” and “Alzheimer’s disease” in all fields. We did not specify article type or language in the initial PubMed search in order to be inclusive of all possibly relevant articles. The search terms were chosen from PubMed MeSH terms that broadly encompassed the intended key topics of CBF and Alzheimer’s disease.

Due to the correlational nature of the research question, the “population, intervention, control, and outcomes” (PICO) method (Richardson et al., 1995) was not used. Unlike an intervention and outcome, CBF and MCI/AD do not have a clear causal relationship. Therefore, the design used for bias and certainty assessments was a case-control research question in which the condition was MCI/AD and the variable of interest was altered CBF.

For the main synthesis, we describe increase or decrease in CBF between AD and controls in several brain regions. Altered CBF in MCI relative to controls and correlations between CBF and cognitive test scores are described as well. For papers that present relevant data only in the form of a graph, we extracted quantitative data using WebPlotDigitizer version 4.5 (automeris.io/WebPlotDigitizer/). For articles that had a high potential of overlapping participant samples, the article with the higher quality rating (less risk for bias) was chosen.

The information recorded for each article were: author(s), article title, year of publication, place of the study, number of participants, diagnostic groups, criteria by which diagnoses were defined, method used to measure and quantify CBF, ages of participants, whether the paper is included in the synthesis, and which outcomes were reported. To organize this information, we used a matrix from University of Maryland Research Guides (lib.guides.umd.edu/SR/steps).

To assess the risk of bias and the quality of each article, we used the NIH Quality Assessment of Case-Control Studies. The items on this tool were: clearly stated objective, clearly specified and defined population, sample size justification, use of same population for cases and controls, use of same inclusion and exclusion criteria for cases and controls, clearly defined and differentiated cases, random selection of sample from eligible individuals, use of concurrent controls, confirmation that exposure occurred prior to the condition, clearly defined and reliable measures of exposure/risk, blinded assessment of exposure/risk with regard to case/control, and use of matching or addition of confounding variables to statistical models. Overall judgments were “poor,” “fair,” or “good.” These decisions were made according to the guidance included in the tool, where it is explained that lack of some criteria correspond to “fatal flaws” while others do not greatly affect the overall quality of the paper. CBF was considered the “exposure,” and because the direction of causation between AD and altered CBF is outside the scope of this review, it was not relevant whether the exposure occurred prior to the condition. Therefore, this question was marked as “not applicable” and not used in the overall bias rating. Other items on the rating tool that were not used were: sample size justification, random selection of sample from eligible individuals, and use of concurrent controls, because these items were all either not present or not reported in all or nearly all papers assessed. In addition, blind assessment of the CBF was not applicable for many of the papers because the CBF was measured by fully automated methods. This item was considered for those papers in which hand-drawn regions of interest were used.

For each relevant paper, we reported mean and SD of CBF in each brain region for each diagnostic group. For articles that used voxel-wise measurement of CBF and reported the z or t scores of peak voxels, these scores, along with cluster sizes and p values are reported. For papers that measured the correlation between CBF and cognitive test scores, r and p values are reported. For papers with mean and SD regional CBF values, effect sizes were calculated, and syntheses were presented for multiple brain regions. To measure effect size, we used Hedge’s g, which is Cohen’s d adjusted for small sample sizes, as well as the 95% confidence interval for Hedge’s g. These effect size measures were chosen because CBF is a continuous measure that is dependent on the methods of scanning and processing, so standardized measures were necessary. Effect size was calculated for the difference in CBF between MCI/AD and control groups (control CBF minus MCI/AD CBF) in the following brain regions: frontal, temporal, parietal, temporoparietal, occipital, posterior cingulate, hippocampus, and thalamus. These regions were chosen because they were the most frequently used across all papers. Thresholds were not employed for the effect sizes; we synthesized results for each brain region across studies by reporting the average, median, and range of effect size scores in each region. All Hedge’s g values and 95% confidence intervals are presented in graphs; positive effect sizes denote relatively decreased CBF in MCI/AD compared to controls. These analyses and graphs were completed using the Effect Size Calculator from the Cambridge Centre for Evaluation and Monitoring (https://www.cem.org/).

A meta-analysis was not performed. Heterogeneity is discussed where there is inconsistency in findings across papers, but statistical analyses regarding heterogeneity were not completed. Sensitivity analyses were also not completed. The scope of this paper is to compile and present the findings in the literature and to discuss overall patterns of results. Reporting bias is taken into account, and certainty ratings for each synthesis are presented in the summary of findings table. Grey literature was not included. Negative findings were included whenever they were reported. For the certainty assessment, we used the GRADE tool from the Cochrane handbook. Scores are “very low,” “low,” “moderate,” and “high.” Since this tool is designed for randomized clinical trials, it instructs the assessor to begin with “moderate” for observational or case-control studies. We rated each synthesis according to the rest of the GRADE criteria but did not strictly begin at “moderate” for each paper because, by design, most of the papers were not clinical trials.

## RESULTS

Of 1197 potential articles resulting from a PubMed search of “brain-blood supply” and “Alzheimer’s disease”, 185 were retained for further assessment and 1012 were excluded based on their abstracts. Of those excluded, 352 were reviews, editorials, commentaries, case studies, and other types of articles that did not match the eligibility criteria. 325 did not use resting CBF as an outcome measure, and 157 used animal models or cell cultures rather than human participants. Finally, 94 did not include participants with various diagnoses or levels of cognitive functioning, and access was not available for 84 articles. Of the 185 retained, 29 articles were ultimately chosen for inclusion in this review, and 156 were excluded. Of these, 97 sampled AD patients who were too young or whose ages were not clearly defined, 25 did not use resting CBF as an outcome, 17 did not compare CBF between diagnostic groups or correlate it with cognitive function, 5 included participants with potentially confounding conditions, 3 were out of scope, 2 included patients with AD who were on psychotropic medication, and 7 were likely to have used the same sample as other articles included in this review. The process of choosing relevant articles is presented in Figure 1.

**Figure 1.**
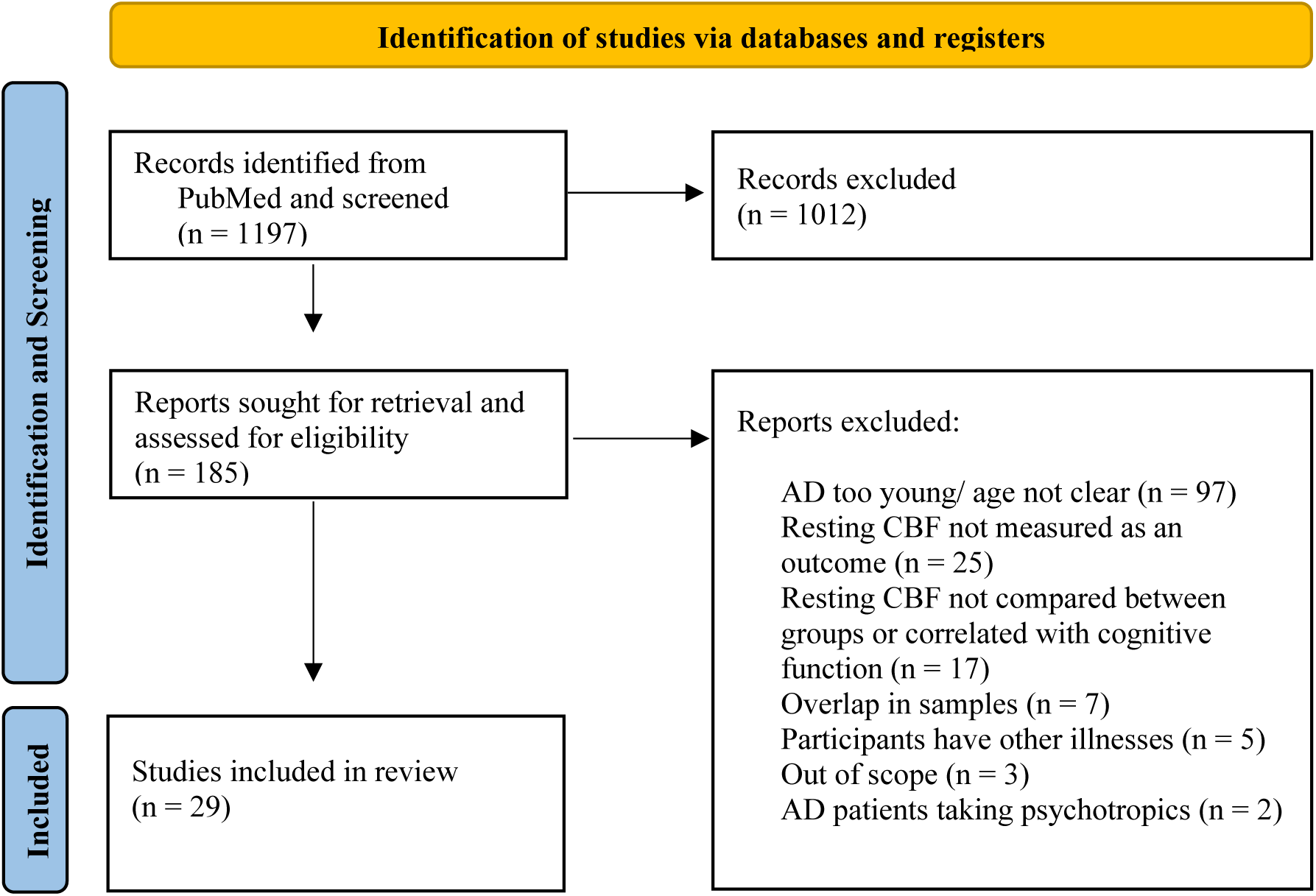
PRISMA flow chart of articles identified, assessed for eligibility, and included in systematic review. Reasons for exclusion are listed. Included papers compare resting CBF between AD/MCI and CN, include an AD sample where over 95% are 60 years old or older, and are written in English.

The papers considered “out of scope” include Iturria-Medina et al. (2017), in which the authors create a 4D multifactorial causal model of AD; Moretti (2016), which focuses on and groups participants by alpha power ratio on EEG; and Vogel et al. (2005), which measures and compares the heterogeneity of CBF rather than a measure of CBF itself. Papers that include patients and/or controls with other illnesses are: Jagust et al. (1990; half of the AD patients have early onset AD), Dougall et al. (2004; some controls have major depression), Starkstein et al. (1994; some controls have dizziness), Cho et al. (2010; controls have headaches or syncope), and De Reuck et al. (1992; the dementia group includes Pick’s disease and Creutzfeldt-Jakob, and the nondemented group includes stroke and encephalopathy). The papers with potentially overlapping samples were: Hanyu et al. (2003; with Hanyu et al. [2004]), Haji et al. (2015; with Kimura et al. [2011]), Brown et al. (1997; with Brown et al. [1996]), Colloby et al. (2002; with Firbank et al. [2011]), Kawamura et al. (1991) and Mortel et al. (1994; with Obara et al. [1994]) and Pearlson et al. (1992; with Harris et al. [1991]).

Papers included in this review are listed and characterized in Supplementary Table 1. They include: Alegret et al. (2010), Benoit et al. (2002), Brown et al. (1996), Claus et al. (1994), Dai et al. (2009), Ding et al. (2014), Encinas et al. (2003), Firbank et al. (2011), Hanyu et al. (2004), Hanyu et al. (2008), Harris et al. (1991), Iizuka and Kameyama (2017), Jagust et al. (1987), Johnson et al. (2007), Kimura et al. (2011), Lacalle-Aurioles et al. (2013), Lee et al. (2003), Mubrin et al. (1989), Nagahama et al. (2003), Obara et al. (1994), Sase et al. (2017), Schuff et al. (2009), Shimizu et al. (2005), Sundstrom et al. (2006), Takahashi et al. (2008), Tateno et al. (2008), van de Haar et al. (2016), Yew and Nation (2017), and Yoshida et al. (2011). Supplementary Table 1 provides the following information for each included article: author, title, year of publication, location of the study (or study authors), sample size by diagnostic group, criteria used to define diagnostic groups, type of scan used to measure CBF, ages of participants in each diagnostic group, whether the paper was included in the synthesis, and which outcomes of interest were included. Notes are included to clarify special circumstances as necessary. Of the 29 papers, 19 measure CBF using SPECT, 6 using ASL MRI, and 4 using other methods. Publication dates range from 1987 to 2017.

Table 1 reports the component scores and overall quality rating on the risk of bias assessment for each paper. Justification for each score is given in the “Overall Quality Rating and Notes” column. Of the 29 papers, 9 were rated as “good,” 13 were rated as “fair,” and 7 were rated as “poor.” All but one of the papers rated “poor” lacked cognitive test scores for the CN group, without which it is not clear that the CN group is free of cognitive decline. Sundstrom et al. (2006) was rated as “poor” because the CN group was younger than the AD group, and age was not used as a covariate in the analyses.

**Table 1.** Risk of bias assessment scores for included articles.

| Authors | Clearly Stated Objective | Clearly Defined Sample | Controls and Cases from same Population | Same Inclusion Criteria for Controls and Cases | Clear Distinction of Cases and Controls | Reliable Methods to Measure CBF | Blind Assessment of CBF | Con-founding Variables | Score and Justification |
| --- | --- | --- | --- | --- | --- | --- | --- | --- | --- |
| Alegret et al. | yes | no | yes | no | yes | yes | not applicable | yes | fair; little information about samples; AD group were all taking AChEIs while the other groups were not, and most CN did not have CT to check for exclusions |
| Benoit et al. | yes | yes | no | not reported | not reported | yes for AD; not reported for CN | not applicable | no | poor; CN are from a different population, scanned separately but “similarly”, not clear whether the inclusion criteria is the same, and cognitive scores for the CN group are not reported |
| Brown et al. | yes | no | yes | yes | yes | yes | yes | no | good |
| Claus et al. | yes | yes | no | yes | yes | yes | yes | yes | good |
| Dai et al. | yes | yes | yes | yes | yes | yes | not applicable | yes | good |
| Ding et al. | yes | yes | not reported | yes | yes | yes | not applicable | no | fair; it is not reported whether the groups are from the same population; only CN are excluded for substance abuse |
| Encinas et al. | yes | yes | yes | yes | yes | yes | yes | no | good |
| Firbank et al. | yes | no | not reported | not reported | yes | yes | yes | no | fair; little information about sample; unclear whether population and/or inclusion criteria are the same between groups |
| Hanyu et al. 2004 | yes | no | not reported | no | yes | yes | not applicable | no | fair; it is not clear whether the groups are from the same population; only AD are excluded based on cerebrovascular lesions |
| Hanyu et al. 2008 | yes | yes for cases, | not reported | not reported | not reported | yes | not applicable | no | poor; essentially no information reported about CN group; |
|  |  | no for controls |  |  |  |  |  |  | unclear whether groups come from the same population or have the same inclusion criteria; no cognitive exam scores reported for CN |
| Harris et al. | yes | yes | yes | yes | yes | yes | yes | no | good |
| Iizuka & Kameyama | yes | yes for cases, no for controls | not reported | not reported | yes | yes | not applicable | no | fair; it is not clear whether the groups are from the same population; only AD are excluded for taking anti-anxiety or antidepressant medications |
| Jagust et al. | yes | no | yes | yes | not reported | yes | not reported | no | poor; while not explicitly clear, it can be deduced that groups are from the same population; inclusion criteria appears to be the same; cognitive exam scores are not reported for CN |
| Johnson et al. | yes | no | yes for the MCI at baseline; unknown for AD | yes | yes | yes | not applicable | no | fair; not clear whether the AD are from the same population as the MCI at baseline. Inclusion criteria is also not applied to several MCI, who are younger than the reported 65 year cutoff |
| Kimura et al. | yes | yes for cases, no for controls | not reported | yes | yes | yes | not applicable | no | fair; little information about the CN group; not clear if they are from the same population as AD |
| Lacalle-Aurioles et al. | yes | yes | yes | yes | yes | yes | yes | no | good |
| Lee et al. | yes | no | not reported | not reported | yes | yes | not applicable | no | fair; little information given about the sample, not clear whether the groups are from the same population or if they have the same inclusion criteria |
| Mubrin et al. | yes | no | not reported | not reported | not reported | yes for imaging; not reported for processing | not reported | no | poor; very little information about samples, especially CN; CN does not have cognitive scores reported |
| Nagahama et al. | yes | yes for cases, no for controls | not reported | yes | not reported | yes | not applicable | no | poor; little information about CN, not sure if they are from the same population as AD; cognitive scores not reported for CN |
| Obara et al. | yes | no | not reported | no | yes | yes | not reported | yes | fair; little information about sample; not sure whether groups are from the same population; inclusion criteria differs slightly between groups |
| Sase et al. | yes | yes | yes | no | yes | yes | not applicable | no | fair; inclusion criteria differs slightly between groups |
| Schuff et al. | yes | yes | yes | yes | yes | yes | not applicable | yes | good |
| Shimizu et al. | yes | yes for cases, no for controls | not reported | no | not reported | yes | not applicable | no | poor; not reported whether the groups are from the same population; only AD are excluded for taking medication that affects the CNS; cognitive exam scores are not reported for CN |
| Sundström et al. | yes | yes | yes | yes | yes | yes | not applicable | no | poor; CN are younger than AD |
| Takahashi et al. | yes | yes | yes | yes | yes | yes | not applicable | no | good |
| Tateno et al. | yes | no | not reported | no | yes | yes | not applicable | no | fair; little information given about the sample, not clear whether the groups are from the same population; only CN have abnormalities on scans as exclusion criteria |
| van de Haar et al. | yes | yes | yes | no | not reported | yes | yes | yes | fair; only AD are excluded for history of stroke; CN are said to complete cognitive exams, but their scores are not reported |
| Yew & Nation | yes | yes | yes | yes | yes | yes | not applicable | yes | good |
| Yoshida et al. | yes | yes | yes | no | yes | yes | yes | no | fair; inclusion criteria are not the same: all AD have *APOE* ε4, but this is not reported for CN |

AD: Alzheimer's disease, AchEI: acetyl cholinesterase inhibitor, *APOE*: apolipoprotein E, CBF: cerebral blood flow, CN: cognitively normal, CNS: central nervous system, CT: computed tomography, MCI: mild cognitive impairment

Table 2 and Supplementary Tables 2 and 3 report the results of interest for each study. Table 2 reports the comparison of CBF in CN and AD groups (notes clarify which papers include MCI in one of these groups), Supplementary Table 2 reports the comparison of CBF between CN and MCI groups, and Supplementary Table 3 presents correlations between CBF and cognitive exam scores. In Table 2 and Supplementary Table 2, regional CBF data is given as mean (SD) for each group. For regional CBF in these tables and further analyses, values for left and right regions were averaged, with SDs pooled; this was also done across smaller brain regions within those of interest. When bilateral averaging was done, or when only one hemisphere was included in a given paper, this is clarified in the “Notes” column. For voxel-wise results, cluster size (k), p value, and z or t value are reported, and the specific region where the peak voxel is located is given in the “Notes” column. For all effect sizes, a positive value represents a *decrease* in CBF in patients relative to CN, as this was how the results were reported in nearly all of the papers. The “Notes” column also mentions if the data were extracted from a graph rather than reported numerically in the paper. Papers with qualitative results are included in this table as well, for completeness. All papers used quantitative or semi-quantitative methods to measure CBF; papers that assessed CBF simply by visual inspection were not included in this review. Supplementary Table 3 reports r and p values for the correlation of regional CBF and cognitive test scores with details in the “Notes” column.

**Table 2.**
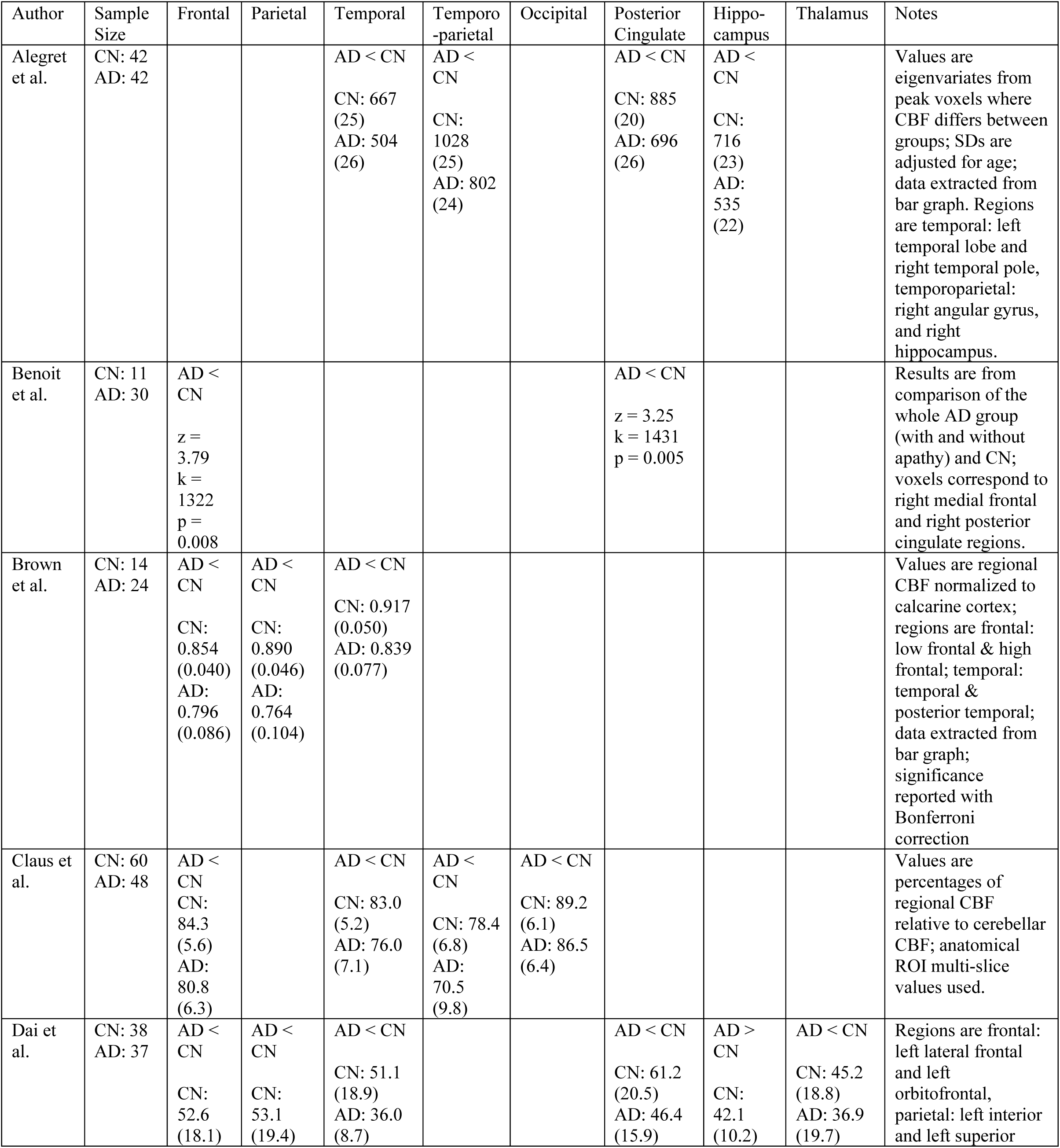
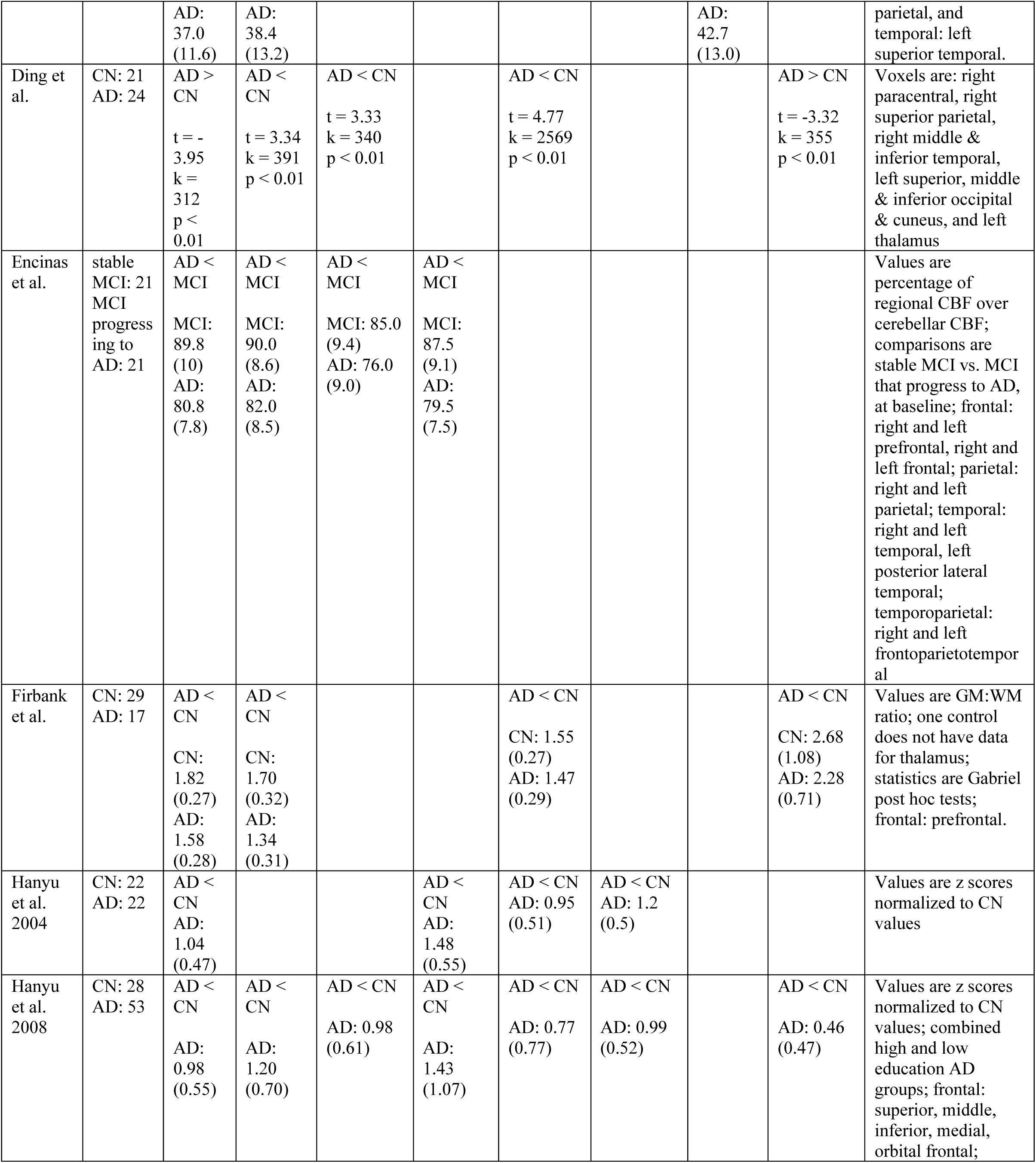
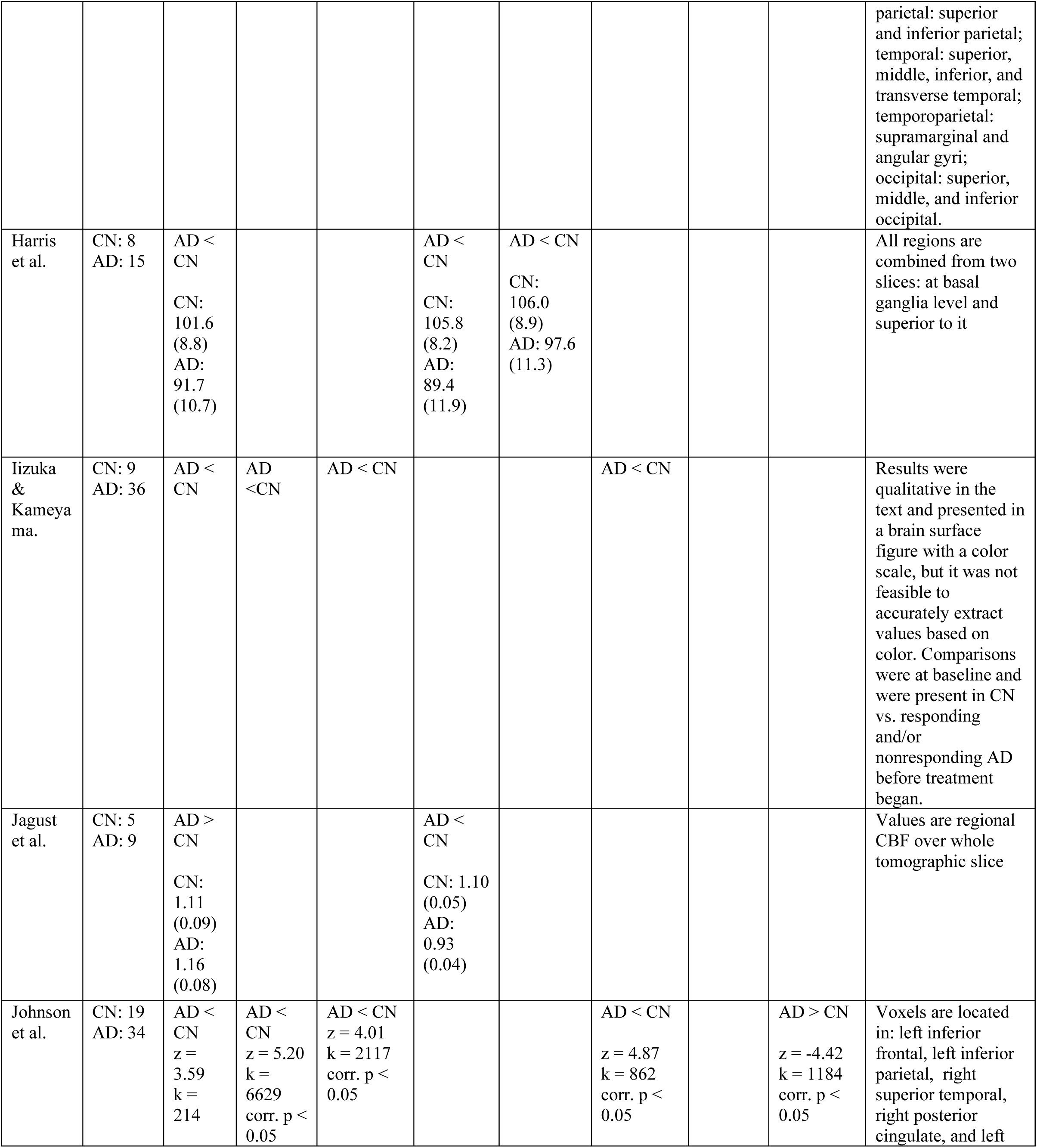
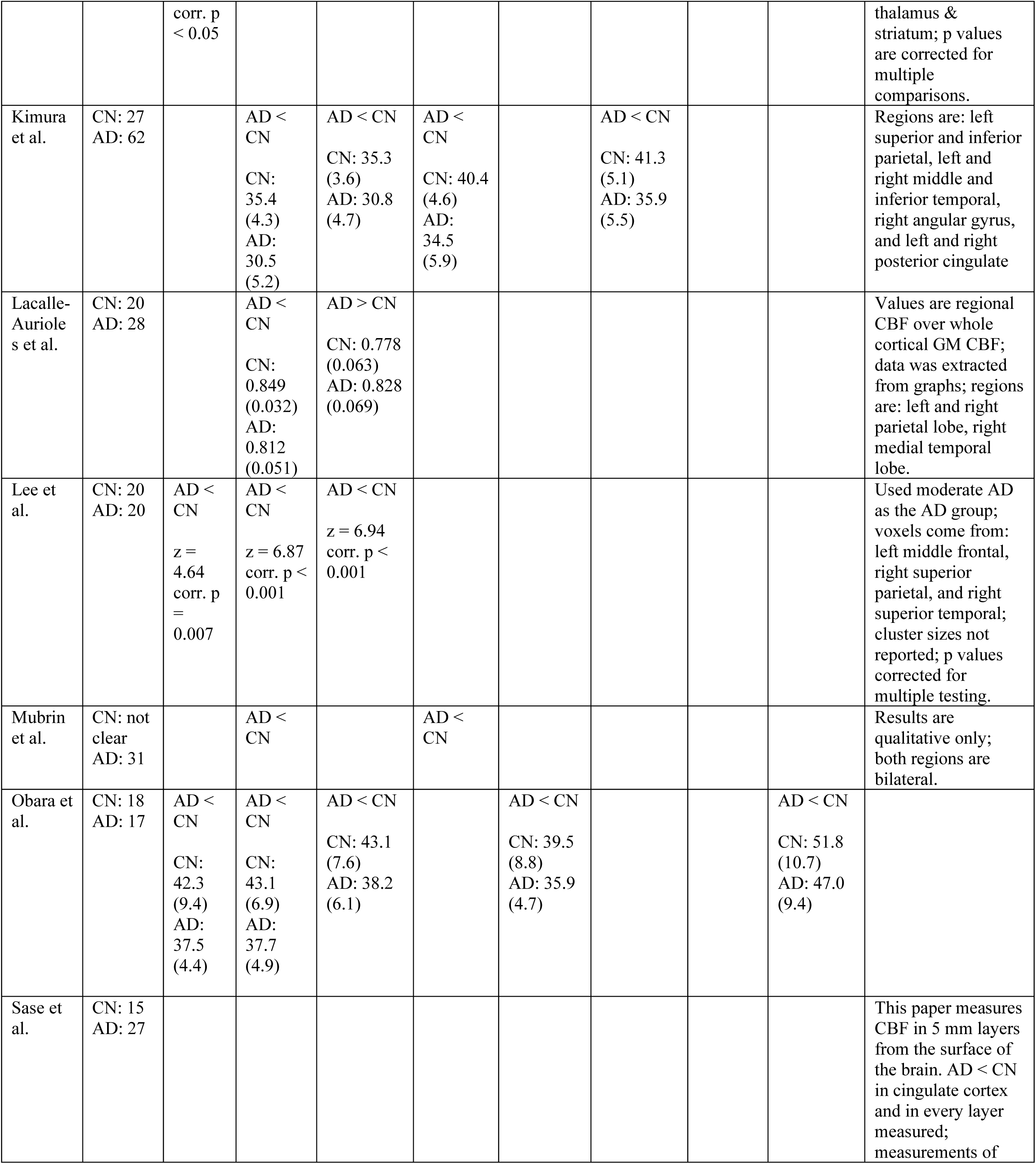
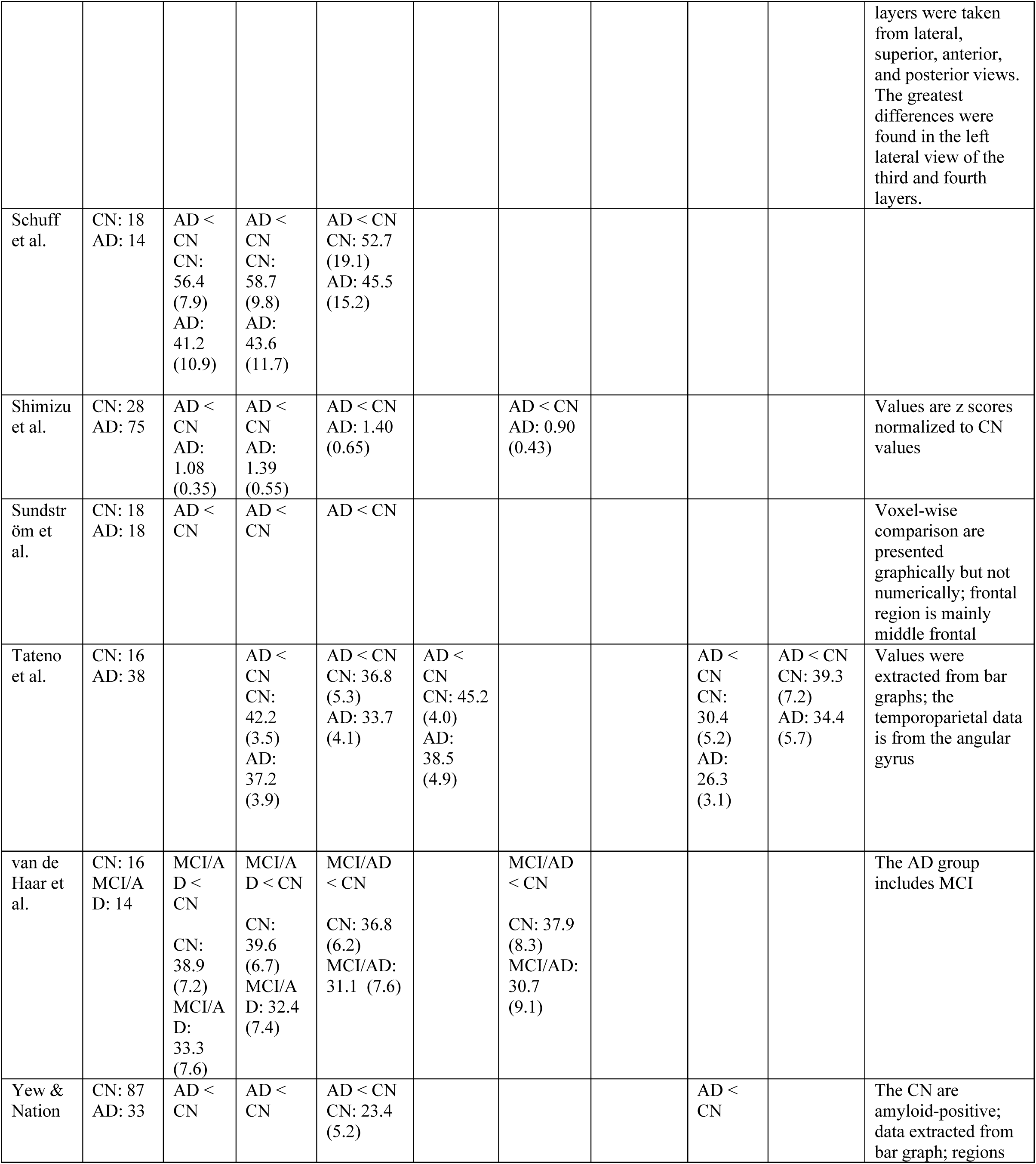
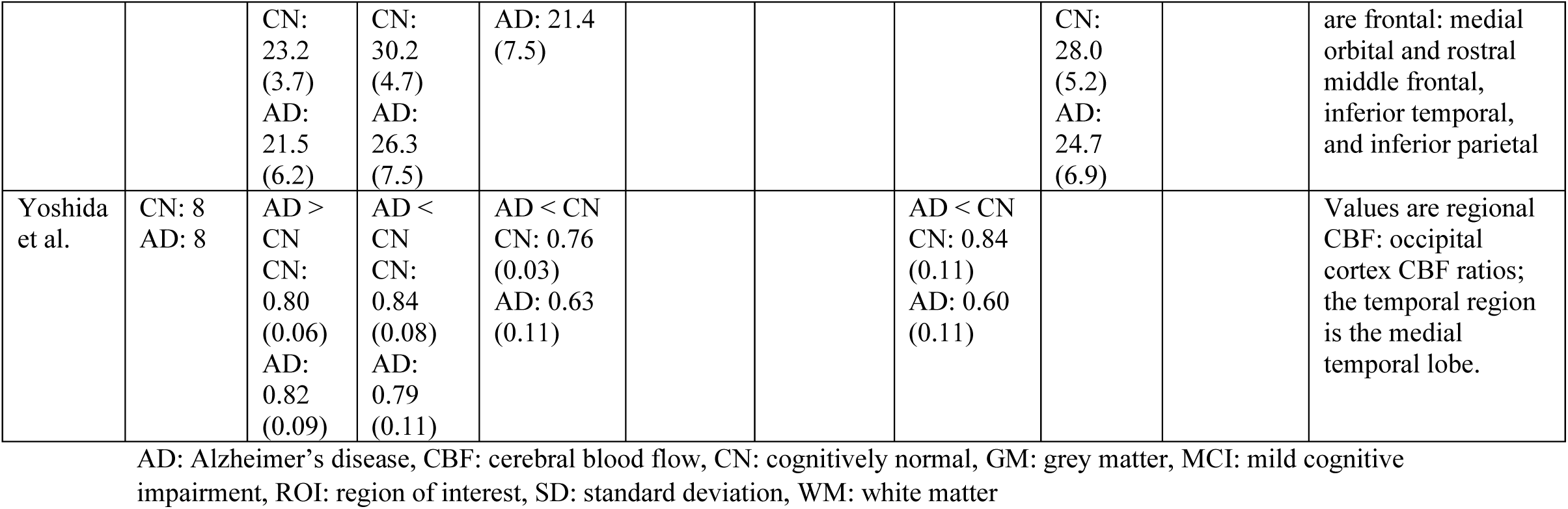
CBF in CN and AD in each paper.

Overall, most of the papers reported decreased CBF in AD relative to controls in all brain regions we included. Increased CBF in AD relative to controls was reported in some papers in the following regions: frontal and temporal lobes, hippocampus, and thalamus. There were fewer papers that assessed CBF in MCI compared to CN, but relatively decreased CBF in MCI was reported in all regions except for the thalamus, and relatively increased CBF in MCI was reported in the frontal and temporal lobes, hippocampus, and thalamus.

In terms of the number of studies that reported each finding, decreased CBF in AD was reported in the frontal lobe in 18 papers, in the parietal lobe in 20 papers, in the temporal lobe in 19 papers, in temporoparietal regions in 10 papers, in the occipital lobe in nine papers, in the posterior cingulate in nine papers, in the hippocampus in three papers, and in the thalamus in five papers. Relative increases in CBF in AD were reported in the frontal lobe in three papers, in the temporal lobe in one paper, in the hippocampus in one paper, and in the thalamus in two papers.

Relative decreases in CBF in MCI were reported in the frontal lobe in one paper, in the parietal lobe in one paper, in the temporal lobe in three papers, in the temporoparietal region in one paper, in the occipital lobe in one paper, in the posterior cingulate in three papers, and in the hippocampus in one paper. Relative increases in CBF in MCI were reported in the parietal lobe in two papers, in the temporal lobe in one paper, in the hippocampus in one paper, and in the thalamus in one paper. These results include reports of altered CBF that were not statistically significant in the original papers.

Of the papers that assessed the relationship between cognitive test scores and CBF, a majority of them reported a positive correlation, but there were also reports of negative correlations and of no correlation. Specifically, a positive correlation between regional CBF and cognitive function was reported in nine papers. The cognitive tests used in these papers include the Mini Mental State Examination, 15-Objects Test, 15 item Boston Naming Test, Poppelreuter test, Cognitive Capacity Screening Examination, Cambridge Cognition Examination (CAMCOG; language, praxis, and abstraction sub-scores), Mini Mental State Questionnaire, Trail Making Test Part B, and the Clock Drawing Test. A negative correlation between regional CBF and scores on the Trail Making Test Part B was reported in one paper. One paper reported no correlation between global CBF and revised CAMCOG scores when other independent variables were in the model, and another paper reported no correlation between regional CBF and the memory sub-score of the CAMCOG. One paper reported no correlation between regional CBF and the California Verbal Learning Test or Self-Ordering Test. Additionally, two papers reported a mix of positive and negative correlations between regional CBF and cognitive test scores.

Figure 2 and Supplementary Figure 1 graphically present the Hedge’s g score and 95% confidence interval for the difference in CBF between AD and CN (Figure 2) and between MCI and CN (Supplementary Figure 1) for each brain region. Positive values represent relatively decreased CBF in MCI/AD. Alegret et al. (2010) reported regional CBF as mean (SD) of eigenvariates from voxel-wise comparisons, which are not directly comparable to the other results, but are included in these graphs for completeness.

**Figure 2.**
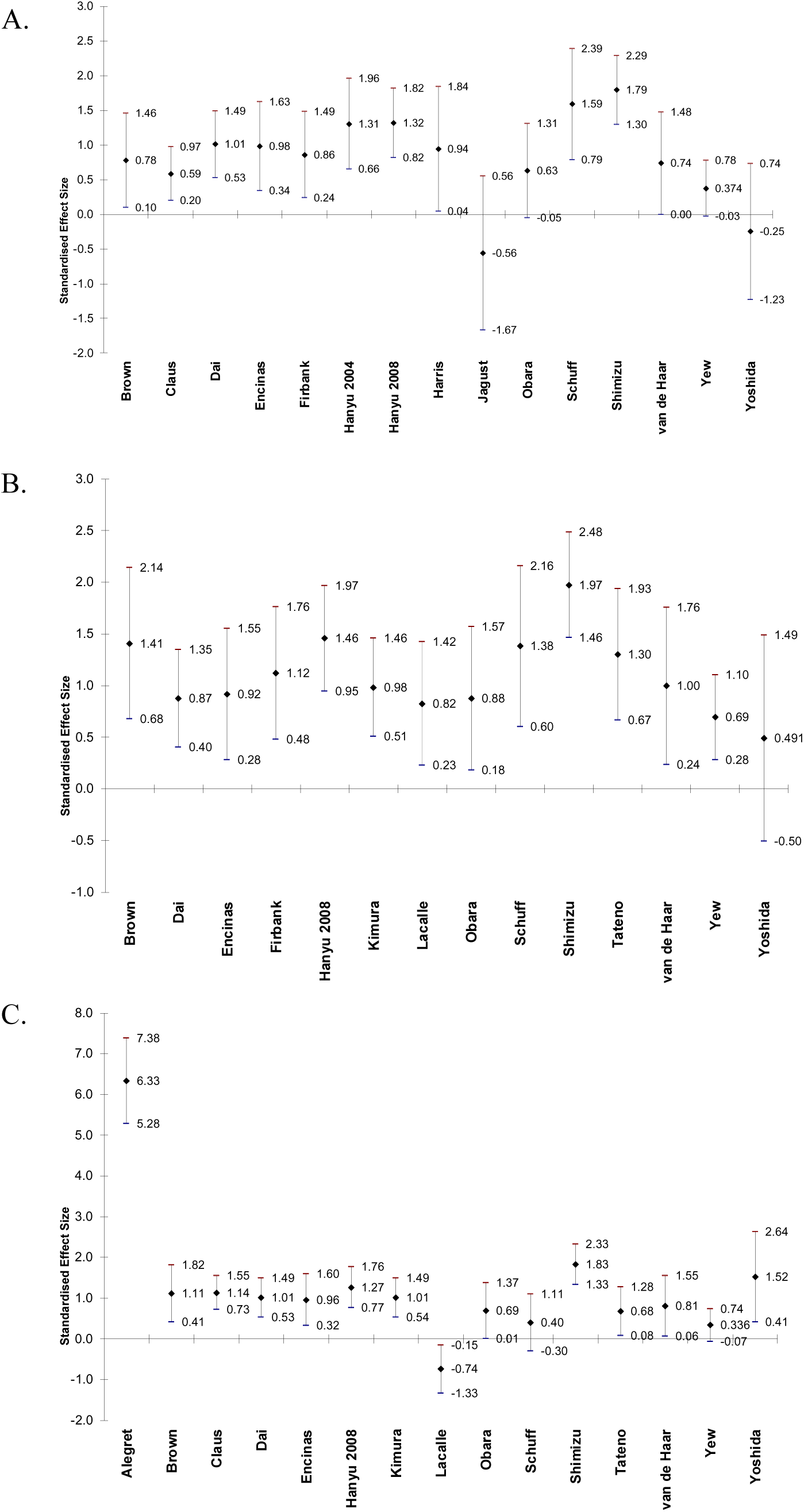
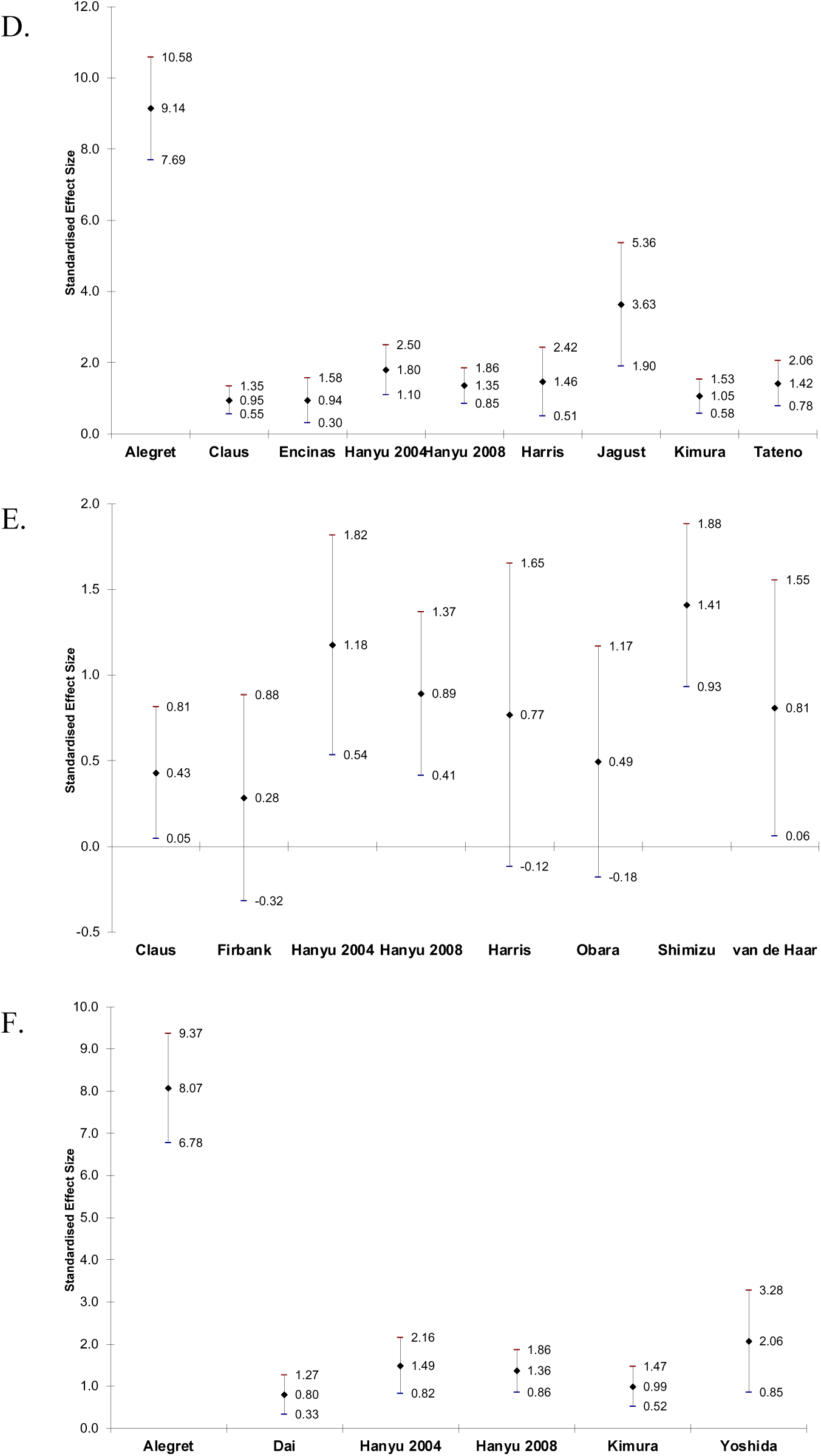
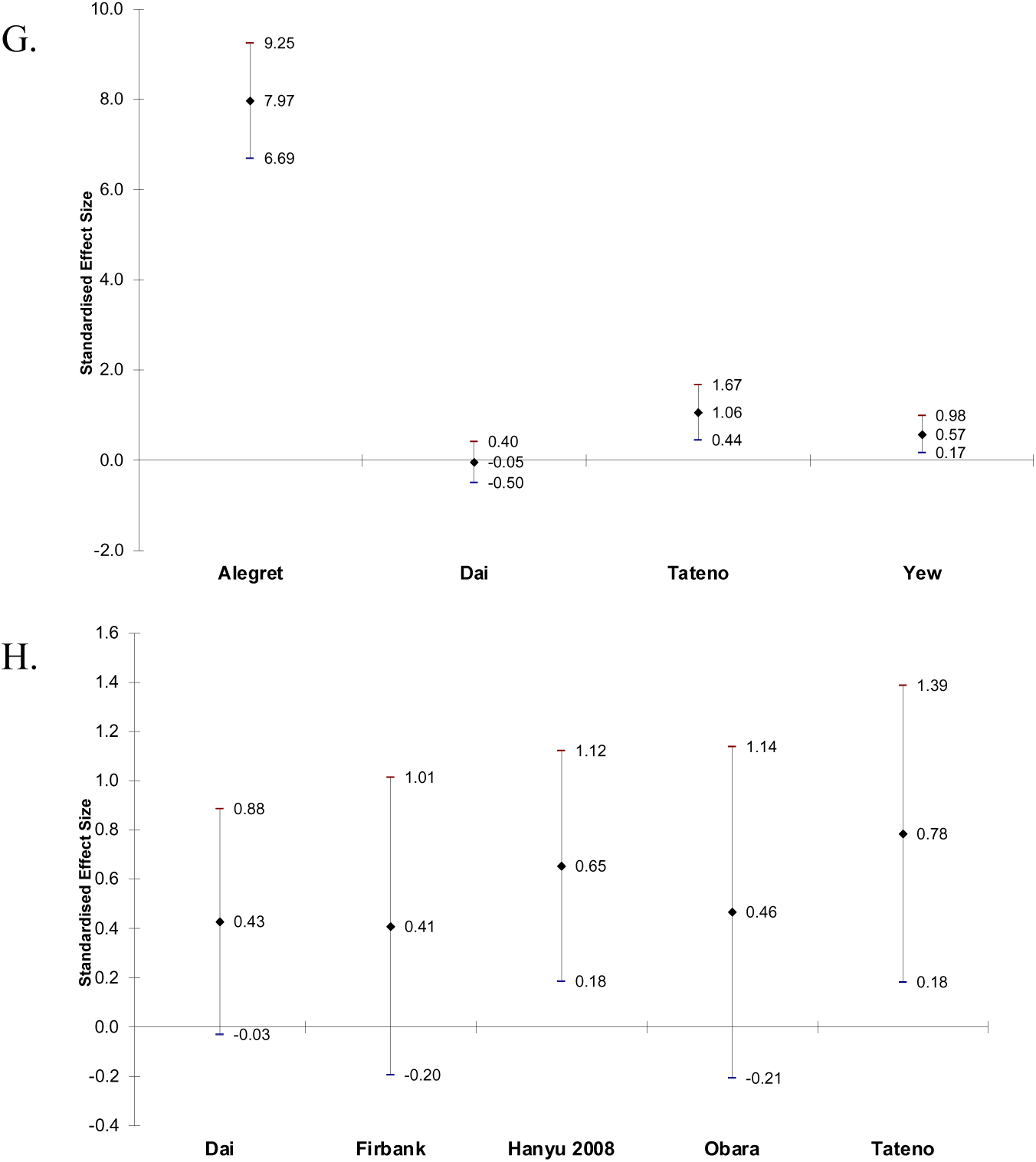
Effect sizes and 95% confidence intervals of difference in CBF between AD and CN in frontal lobe (A), parietal lobe (B), temporal lobe (C), temporoparietal region (D), occipital lobe (E), posterior cingulate (F), hippocampus (G), and thalamus (H). Positive effect sizes indicate that CBF is decreased in AD relative to CN.

Table 3 is a Summary of Findings table for the syntheses of results. Results are presented for CBF in CN vs. AD in papers that reported regional CBF values as mean (SD). Seventeen papers were included. Alegret et al. (2010) was not included. Encinas et al. (2003) was also not included because there was no CN group in that study. Table 3 reports combined sample size, number of studies included, authors of included studies, mean, median, and range of Hedge’s g scores, certainty, and comments for syntheses of CBF in AD relative to controls in each brain region. The region with the greatest mean Hedge’s g (greatest relative decrease in CBF in AD) is the temporoparietal (g = 1.67), followed by the posterior cingulate (g = 1.34). All regions had positive mean and median Hedge’s g scores (decreased relative CBF in AD). At least one paper reported an increase in CBF in AD compared to CN in the frontal lobe, temporal lobe, and the hippocampus.

**Table 3.** Syntheses of CBF in AD compared to CN by brain region.

| Brain Region | Individuals | Studies | Hedge's G; Mean, Median and Range | Certainty | Comments |
| --- | --- | --- | --- | --- | --- |
| Frontal | 386 cases;<br>379 controls | 14: Brown et al., Claus et al., Dai et al., Firbank et al., Hanyu et al. 2004, Hanyu et al. 2008, Harris et al., Jagust et al., Obara et al., Schuff et al., Shimizu et al., van de Haar et al., Yew and Nation, Yoshida et al. | 0.79;<br>0.82<br>(-0.56, 1.79) | low | Certainty lowered because: data extracted from graphs (Brown et al., Jagust et al., Yew and Nation), several subregions and high and low education groups combined in Hanyu et al. 2008, subregions combined in Brown et al., Harris et al.; Notes: two papers with increase in AD (Jagust et al. and Yoshida et al.) have very small sample sizes, van de Haar et al. has MCI in AD group and Yew and Nation has MCI in CN group. All AD in Yoshida et al. are *APOE* ε4 positive. |
| Parietal | 420 cases;<br>347 controls | 13: Brown et al., Dai et al., Firbank et al., Hanyu et al. 2008, Kimura et al., Lacalle-Aurioles et al., Obara et al., Schuff et al., Shimizu et al., Tateno et al., van de Haar et al., Yew and Nation, Yoshida et al. | 1.11;<br>1.00<br>(0.49, 1.97) | moderate | Certainty lowered because: data extracted from graphs (Brown et al., Lacalle-Aurioles et al., Tateno et al., Yew and Nation), several subregions and high and low education groups combined in Hanyu et al. 2008, subregions combined in Brown et al. Notes: van de Haar et al. has MCI in AD group and Yew and Nation has MCI in CN group, all AD in Yoshida et al. are *APOE* ε4 positive. |
| Temporal | 451 cases;<br>378 controls | 13: Brown et al., Claus et al., Dai et al., Hanyu et al. 2008, Kimura et al., Lacalle-Aurioles et al., Obara et al., Schuff et al., Shimizu et al., Tateno et al., van de Haar et al., Yew and Nation, Yoshida et al. | 0.85;<br>1.01<br>(-0.74, 1.83) | moderate | Certainty lowered because: data extracted from graphs (Brown et al., Lacalle-Aurioles et al., Tateno et al., Yew and Nation), several subregions and high and low education groups combined in Hanyu et al. 2008, subregions combined in Brown et al.; Notes: one paper with increase in AD (Lacalle-Aurioles et al.) shows same pattern to a lesser extent in MCI; van de Haar et al. has MCI in AD group and Yew and Nation has MCI in CN group, all AD in Yoshida et al. are *APOE* ε4 positive. |
| Temporoparietal | 247 cases;<br>166 controls | 7: Claus et al., Hanyu et al. 2004, Hanyu et al. 2008, Harris | 1.67;<br>1.42<br>(0.95, 3.63) | moderate | Certainty lowered because: data extracted from graphs (Jagust et al., Tateno et al.), several subregions and high and low education groups |
|  |  | et al., Jagust et al., Kimura et al., Tateno et al. |  |  | combined in Hanyu et al. 2008, subregions combined in Harris et al.; Notes: Jagust et al. had the largest effect size despite a small sample size, and this paper also showed a positive correlation between CBF and cognitive scores. |
| Occipital | 261 cases;<br>209 controls | 8: Claus et al., Firbank et al., Hanyu et al. 2004, Hanyu et al. 2008, Harris et al., Obara et al., Shimizu et al., van de Haar et al. | 0.78;<br>0.79<br>(0.28,<br>1.41) | low | Certainty lowered due to low cumulative bias rating of constituent papers, several subregions and high and low education groups combined in Hanyu et al. 2008, subregions combined in Harris et al.; Notes: van de Haar et al. has MCI in AD group. |
| Posterior Cingulate | 182 cases;<br>123 controls | 5: Dai et al., Hanyu et al. 2004, Hanyu et al. 2008, Kimura et al., Yoshida et al. | 1.34;<br>1.36<br>(0.80,<br>2.06) | low | Certainty lowered due to low cumulative bias rating of constituent papers, several subregions and high and low education groups combined in Hanyu et al. 2008; Notes: all AD in Yoshida et al. are *APOE* ε4 positive. |
| Hippocampus | 108 cases;<br>141 controls | 3: Dai et al., Tateno et al., Yew and Nation | 0.53;<br>0.57<br>(-0.05,<br>1.06) | low | Certainty lowered because: data extracted from graphs (Tateno et al., Yew and Nation); Notes: one paper with increase in AD (Dai et al.) shows similar pattern in MCI, but the effect is very small in AD. Yew and Nation has MCI in CN group.. |
| Thalamus | 162 cases;<br>129 controls | 5: Dai et al., Firbank et al., Hanyu et al. 2008, Obara et al., Tateno et al. | 0.55;<br>0.46<br>(0.41,<br>0.78) | moderate | Certainty lowered due to low cumulative bias rating of constituent papers, data extracted from graphs (Tateno et al.), several subregions and high and low education groups combined in Hanyu et al. 2008; Notes: Firbank et al. is missing this region for one control. |

AD: Alzheimer's disease, *APOE*: apolipoprotein E, CBF: cerebral blood flow, CN: cognitively normal, MCI: mild cognitive impairment

Reporting bias was assessed by rating the completeness of each paper in the syntheses. All papers that reported mean and SD CBF values for AD and CN were included except for Alegret et al. (2010) due to this paper’s use of voxel-wise eigenvariates. Of the included papers, Firbank et al. (2011) was missing CBF data from the thalamus for one CN participant, and Schuff et al. (2009) did not analyze or report occipital CBF data because too few voxels were present after selecting only “pure” gray or white matter voxels. Otherwise, results were fully reported in all papers. Papers were also included in regional syntheses if they reported CBF values for relevant subregions, some of which we combined for an average value within the synthesis region. This review only includes published articles from PubMed that were in English and fully accessible. These restrictions contribute to a moderate level of reporting bias, but reporting bias is likely not increased from other sources or for other reasons.

Certainty of syntheses was rated based on cumulative risk of bias scores of the constituent papers; if the weighted average risk of bias score (based on sample size) was less than “fair,” the overall certainty was lowered. Certainty was also lowered due to inconsistency (scores were higher if all results were in the same direction), and indirectness and/or imprecision (including papers where we combined subregions’ values, subgroups, or where we extracted data from graphs). Large effect sizes, evidence of dose-response relationships (MCI values intermediate between CN and AD values), and confounding factors that would have likely resulted in an effect of the opposite direction were considered as reasons to raise or maintain the certainty score. Justifications for the certainty score are included in detail in Table 3 for each synthesis.

## DISCUSSION

In this systematic review, we compiled findings of altered CBF in MCI/AD from previously published literature. There were consistent reports of decreased CBF in AD compared to CN in parietal, temporoparietal, occipital, and posterior cingulate regions. Frontal, temporal, hippocampal, and thalamic regions each had either increases in CBF in AD or no differences reported in at least one paper. Interestingly, thalamic CBF was relatively decreased in AD in all five papers that reported regional CBF and relatively increased in the two papers that reported voxel-wise results. The greatest relative decrease in CBF in AD was in the temporoparietal and posterior cingulate regions. Across regions, 93% of the results reported decreased CBF in AD compared to CN. For CBF in MCI compared to CN, the results were mixed. Only five papers reported regional or voxel-wise comparisons in CBF between CN and MCI, but of the 16 regional results, 5 (31%) of them reported increased CBF in MCI compared to CN. The higher percentage of brain regions with relatively increased CBF in MCI may be because there were fewer papers that compared CBF between MCI and CN. However, it has been suggested that increased CBF in MCI could be a compensatory response to early AD pathology, in which increased blood flow is necessary in order to function at a normal level. In Zhang et al. (2021), the authors concluded that decreased CBF begins in the posterior cingulate, temporoparietal, and thalamic regions during MCI and that decreased CBF becomes more widespread in AD. We found some evidence of decreased CBF in MCI and reported MCI values that were intermediate between CN and AD values (Alegret et al., 2010; Dai et al., 2009). We also found that the greatest relative decreases in CBF in AD were located in the temporoparietal region and the posterior cingulate. Both of these regions have been reported to have decreased CBF and decreased glucose metabolism in previous studies and reviews, and these alterations are considered characteristic of AD (Matsuda, 2001). The reasons for this pattern may be due to the spatial spread of tau pathology during AD, to vulnerability of these areas to vascular and/ or neuronal damage, or to damage of functionally connected brain regions. Like Zhang et al. (2021), we found quite consistent findings of positive correlations between regional CBF and cognitive functioning, which supports the idea that decreased CBF is associated with cognitive impairment in AD. It will be necessary for future studies to elucidate whether transient increases in CBF during MCI are characteristic of this stage of disease or if this only occurs in a particular subset of individuals with MCI.

Another finding from this review is that only a few papers comparing CBF between diagnostic stages accounted for potential confounding variables other than age in the analyses. Nearly all papers accounted for the potential effect of age on CBF by using diagnostic groups that did not differ by age. Sundstrom et al. (2006) reported that the CN group was younger than the AD group. The only paper that did not report adequate information about group age was Mubrin et al. (1989). Neither of these papers were included in the syntheses presented in the Summary of Findings Table. Aside from age, three papers included sex as a covariate, one paper included the presence of hypertension, one paper included white matter lesion volume, one paper included the presence of the *APOE* ε4 allele, one paper included body mass index, and one paper included cerebral glucose metabolism. These factors and others, including other cardiovascular risk factors, lifestyle factors, and possibly socioeconomic factors could potentially have an effect on CBF and/or cognitive function, and they should be taken into account in future studies of CBF in AD.

One of the limitations of this review is that we included only accessible articles from PubMed that were published in English. Another limitation inherent to this review is the inclusion of articles across various decades, in which several different imaging and processing methods were used to measure CBF. Papers also used different reference regions to normalize the CBF values. Despite this, the results of CBF in AD compared to CN and of CBF in relation to cognitive test scores were generally consistent. The less consistent results in the MCI group may be due to the small sample size or may be due to differences that were not accounted for in the MCI samples. Additionally, this review only includes papers with AD samples in a specific age range, so the number of papers included is relatively small, and some of them have small sample sizes. Due to these limitations and to the observational and correlational nature of the research question, the results and syntheses presented here are meant to summarize and consolidate the existing evidence of altered CBF in MCI/AD and to characterize general patterns in these findings, rather than to make conclusive statements about the degree of these changes.

## CONCLUSIONS

The results of this review suggest that more research is needed to better understand the spatial and temporal nature of changes in CBF throughout the course of AD, especially in early stages of disease including MCI. In addition, standardization of methods to measure and process CBF would allow results from different studies to be more easily consolidated or compared. We believe that to best characterize CBF for its potential use as an early noninvasive biomarker for AD, large and diverse samples of participants must be employed so that analyses can be completed in multiple brain regions and with multiple factors included as covariates. Longitudinal studies that meet these criteria and include individuals from different disease stages will likely help to clarify the specific patterns of altered CBF that are usually present during the years leading up to a diagnosis of AD, as well as determine how alterations in CBF relate to AD pathologies and cognitive decline.

## Author Contributions

Author contributions included conceptual and study design (CGS, SLR and AJS), literature search and selection of included publications (CGS), summarization, synthesis, and presentation of results (CGS), interpretations, conclusions, and overall messaging (all authors), and comments, revisions, and approval for final publication (all authors).

## Data Availability Statement

We did not analyze or generate datasets because this work involved the compilation and synthesis of previously published results, all of which are cited and available to view.

## Supporting information

PRISMA Checklist

---

**Supplementary Figure 1.**
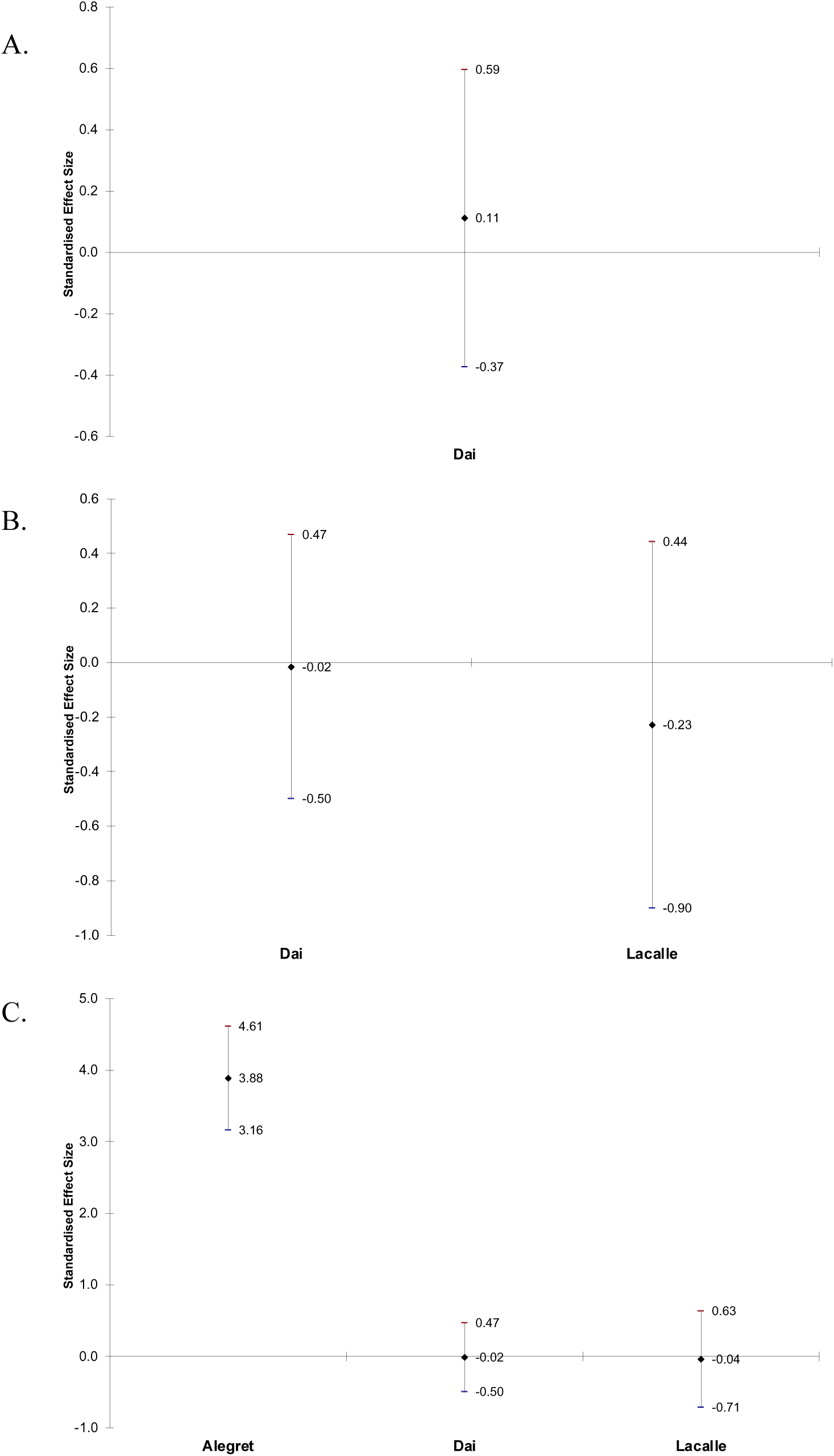
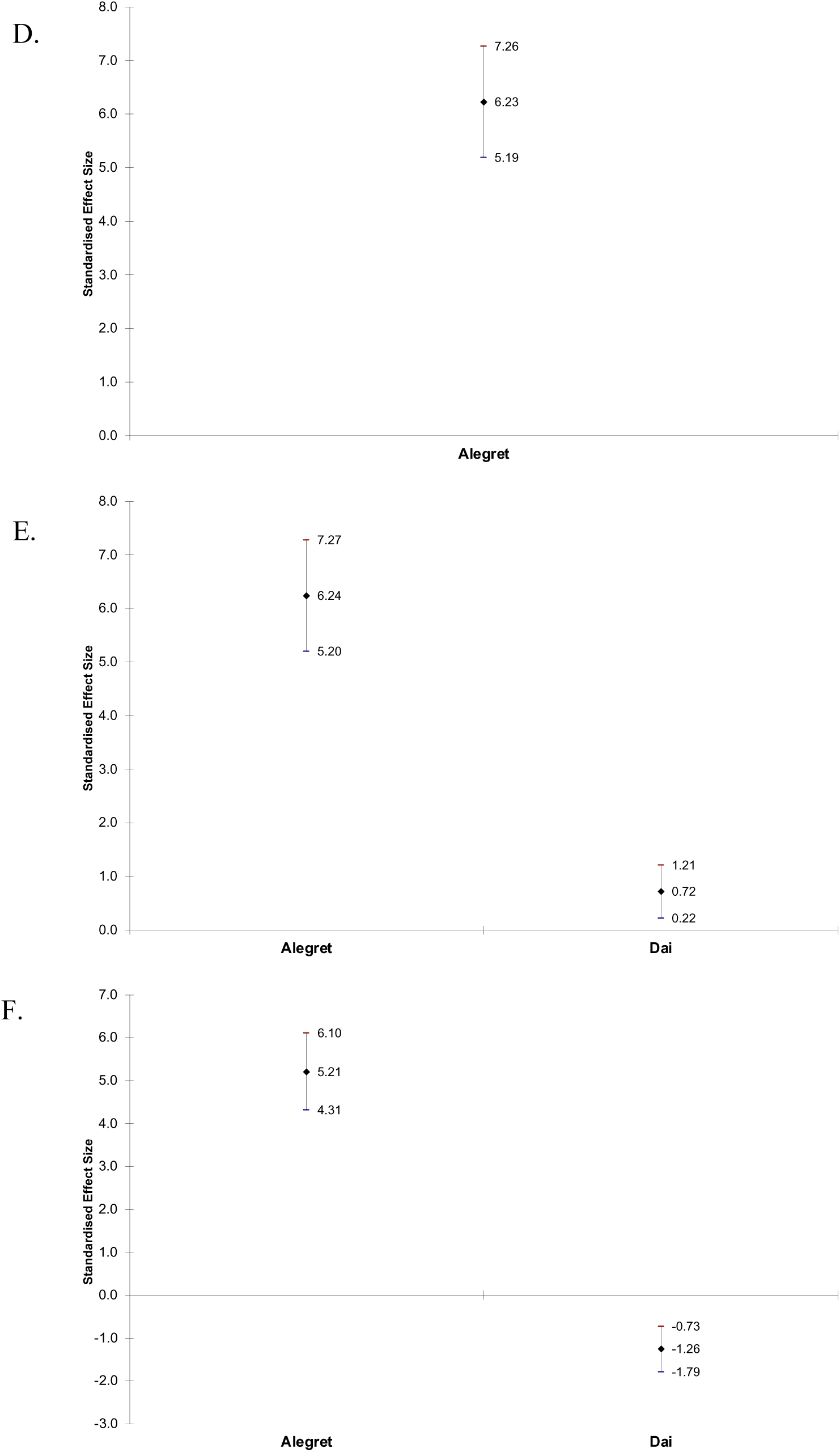
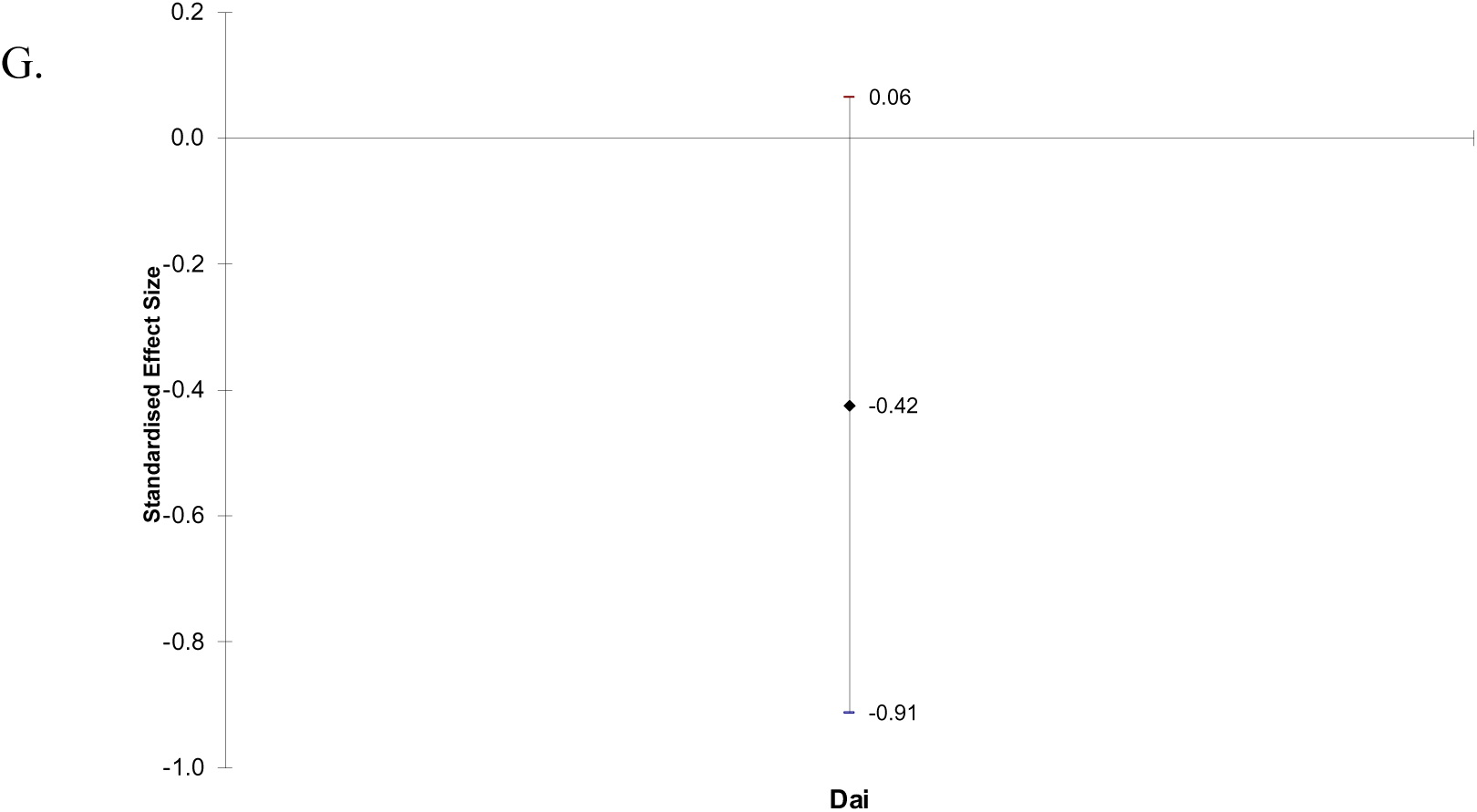
Effect sizes and 95% confidence intervals of difference in CBF between MCI and CN in frontal lobe (A), parietal lobe (B), temporal lobe (C), temporoparietal region (D), posterior cingulate (E), hippocampus (F), and thalamus (G). Positive effect sizes indicate that CBF is decreased in MCI relative to CN.

**Supplementary Table 1.** Characteristics of included articles.

| Authors | Title | Publication Date | Location of Study/ Authors | Number of Participants and Diagnostic Groups | Criteria for Diagnoses | CBF Measurement Method | Ages by Diagnostic Group (mean [SD]) | Results Included in Syntheses | Reports Relative CBF for AD | Reports Relative CBF for MCI | Reports Association between CBF and Cognitive Test Scores |
| --- | --- | --- | --- | --- | --- | --- | --- | --- | --- | --- | --- |
| Alegret et al. | Brain perfusion correlates of visuoperceptual deficits in mild cognitive impairment and mild Alzheimer's disease | 2010 | Barcelona, Spain | 126 (42 CN, 42 MCI, 42 AD) | NINCD S-ADRD A for AD and Petersen et al criteria for MCI | <sup>99m</sup>Tc-ECD SPECT | CN: 74.7 (4.4)<br>MCI: 76.8 (4.3)<br>AD: 76.4 (4.5) | no | yes | yes | yes |
| Benoit et al. | Brain perfusion in Alzheimer's disease with and without apathy: a SPECT study with statistical parametric mapping analysis | 2002 | Nice University Memory Center, France | 41 (11 CN, 30 AD) | ICD-10 criteria | <sup>99m</sup>Tc-ECD SPECT | CN: 74.1 (9.3)<br>AD: 77.1 (5.6) | no | yes | no | yes |
| Brown et al. | Longitudinal changes in cognitive function and regional cerebral function in Alzheimer's disease: a SPECT blood flow study | 1996 | Glasgow, Scotland | 38 (14 CN, 24 AD) | CAMD EX and DSM-III-R | <sup>99m</sup>Tc-HMPAO SPECT | CN: 72 (2.3)<br>AD: 76 (1.5)<br>comparison not described | yes | yes | no | yes |
| Claus et al. | Assessment of cerebral perfusion with single-photon emission tomography in normal subjects and in patients with | 1994 | Netherlands; Rotterdam Elderly Study | 108 (60 CN, 48 AD) | NINCD S-ADRD A | <sup>99m</sup>Tc-HMPAO SPET | CN: 74.1 (0.8)<br>MCI: 72.2 (1.2) | yes | yes | no | no |
|  | Alzheimer's disease: effects of region of interest selection |  |  |  |  |  |  |  |  |  |  |
| Dai et al. | Mild cognitive impairment and alzheimer disease: patterns of altered cerebral blood flow at MR imaging | 2009 | Pittsburgh, USA; Cardiovascular Health Study Cognition Study | 104 (38 CN, 29 MCI, 37 AD) | described in Lopez et al 2003 | CASL MRI | CN: 82.1 (3.6)<br>AD: 83.6 (3.5) | yes | yes | yes | no |
| Ding et al. | Pattern of cerebral hyperperfusion in Alzheimer's disease and amnesic mild cognitive impairment using voxel-based analysis of 3D arterial spin-labeling imaging: initial experience | 2014 | China | 62 (21 CN, 17 MCI, 24 AD) | NINCDS-ADRDA for AD and Petersen et al criteria for MCI | 3D pCASL MRI | CN: 69.6 (5.9)<br>MCI: 71.4 (7.6)<br>AD: 74.6 (6.7) | no | yes | yes | no |
| Encinas et al. | Regional cerebral blood flow assessed with <sup>99m</sup>Tc-ECD SPET as a marker of progression of mild cognitive impairment to Alzheimer's disease | 2003 | Madrid, Spain | 42 (21 MCI, 21 AD)<br>all start with MCI and some progress to AD | International Psychogeriatric Association and the Alzheimer's Disease Cooperative Study, Global Deterioration Scale (GDS) for | <sup>99m</sup>Tc-ECD SPET | MCI: 71.6 (8.2)<br>AD: 75.3 (5.5) | no | yes | yes | no |
|  |  |  |  |  | MCI;<br>NINCD<br>S-ADRD<br>A,<br>DSM-IV,<br>GDS<br>for AD |  |  |  |  |  |  |
| Firbank<br>et al. | Cerebral<br>blood flow<br>by arterial<br>spin<br>labeling in<br>poststroke<br>dementia | 2011 | Newcas<br>tle upon<br>Tyne,<br>UK | 46 (29<br>CN, 17<br>AD)<br>also a<br>post-<br>stroke<br>group | NINCD<br>S-ADRD<br>A | ASL<br>MRI | CN: 82.9<br>(3.5)<br>AD: 83.2<br>(3.7) | yes | yes | no | yes |
| Hanyu<br>et al.<br>2004 | Cerebral<br>blood flow<br>patterns in<br>Binswanger<br>'s disease: a<br>SPECT<br>study using<br>three-<br>dimensional<br>stereotactic<br>surface<br>projections | 2004 | Tokyo,<br>Japan | 44 (22<br>CN, 22<br>AD)<br>also a<br>Binswang<br>er's<br>disease<br>group | NINCD<br>S-ADRD<br>A | <sup>123</sup>I-IMP<br>SPEC<br>T | CN: 77.1<br>(5.8)<br>AD: 77.9<br>(5.5) | yes | yes | no | no |
| Hanyu<br>et al.<br>2008 | The effect<br>of<br>education<br>on rCBF<br>changes in<br>Alzheimer's<br>disease: a<br>longitudinal<br>SPECT<br>study | 2008 | Tokyo,<br>Japan | 81 (28<br>CN, 53<br>AD)<br>AD<br>grouped<br>by level of<br>education,<br>cutoff 12<br>years | NINCD<br>S-ADRD<br>A | <sup>123</sup>I-IMP<br>SPEC<br>T | CN: 75.1<br>(6.4)<br>AD high<br>education:<br>76.8 (5.7)<br>AD low<br>education:<br>75.9 (6.4) | yes | yes | no | no |
| Harris et<br>al. | Cortical<br>circumferen<br>tial profile<br>of SPECT<br>cerebral<br>perfusion in<br>Alzheimer's<br>disease | 1991 | Baltimo<br>re,<br>Marylan<br>d, USA | 23 (8 CN,<br>15 AD) | NINCD<br>S-ADRD<br>A | <sup>123</sup>I-IMP<br>SPEC<br>T | CN: 70.1<br>(4.9)<br>AD: 72.6<br>(5.9) | yes | yes | no | no |
| Iizuka &<br>Kameyam<br>a | Cholinergic<br>enhanceme<br>nt increases<br>regional<br>cerebral<br>blood flow<br>to the<br>posterior<br>cingulate | 2017 | Tokyo,<br>Japan | 45 (9 CN,<br>36 AD)<br>AD<br>grouped<br>by<br>response<br>to<br>treatment; | NINCD<br>S-ADRD<br>A,<br>DSM-IV-R,<br>and<br>ICD-10 | <sup>123</sup>I-IMP<br>SPEC<br>T | CN: 77.11<br>(3.6)<br>AD: 77.6,<br>range 69-89 | no | yes | no | no |
|  | cortex in mild Alzheimer's disease |  |  | used data from before treatment |  |  |  |  |  |  |  |
| Jagust et al. | The diagnosis of dementia with single photon emission computed tomography | 1987 | California, USA | 14 (5 CN, 9 AD)<br>also 2 with multi-infarct dementia | NINCD S-ADRD A and DSM-III | <sup>123</sup>I-IMP SPEC T | CN: 70 (6)<br>AD: 71 (5) | yes | yes | no | yes |
| Johnson et al. | Single photon emission computed tomography perfusion differences in mild cognitive impairment | 2007 | Massachusetts, USA | 158 (19 CN, 105 MCI, 34 AD) | NINCD S-ADRD A for AD; Clinical Dementia Rating Sum of Boxes for grouping at follow-up | <sup>99m</sup>Tc-HMPAO SPEC T | CN: 73.1 (3.6)<br>MCI: 73.7 (5.0)<br>AD: 75.1 (3.9) | no | yes | yes | yes |
| Kimura et al. | Relationship between thyroid hormone levels and regional cerebral blood flow in Alzheimer disease | 2011 | Oita, Japan | 89 (27 CN, 62 AD) | NINCD S-ADRD A | <sup>99m</sup>Tc-ECD SPEC T | CN: 75.8 (6.5)<br>AD: 77.3 (6.9) | yes | yes | no | no |
| Lacalle-Auriolles et al. | Is the cerebellum the optimal reference region for intensity normalization of perfusion MR studies in early Alzheimer's disease? | 2013 | Madrid, Spain | 63 (20 CN, 15 MCI, 28 AD) | Winblad criteria for MCI, NINCD S-ADRD A for AD | echo planar MRI for perfusion weighted imaging | CN: 71.7 (7.0)<br>MCI: 70.9 (9.7)<br>AD: 74.7 (7.0) | yes | yes | yes | no |
| Lee et al. | Statistical parametric mapping of brain | 2003 | Taiwan | 60 (20 CN, 20 mild AD, 20 | NINCD S-ADRD A for | <sup>99m</sup>Tc-HMPAO | CN: 73.8 (2.8)<br>mild AD: 74.2 (3.0) | no | yes | no | no |
|  | SPECT perfusion abnormalities in patients with Alzheimer's disease |  |  | moderate AD) | AD, CDR, MMSE and Cognitive Abilities Screening Instruments (CASI) for AD severity | SPECT | moderate AD: 74.1 (3.2) |  |  |  |  |
| Mubrin et al. | Normalization of rCBF pattern in senile dementia of the Alzheimer's type | 1989 | Yugoslavia | 26 (CN and AD)<br>numbers per group unknown | DSM-III and elimination of alternative diagnoses | <sup>133</sup>Xenon CT | full sample: 76.2 (8.0) | no | yes | no | no |
| Nagahama et al. | Cerebral correlates of the progression rate of the cognitive decline in probable Alzheimer's disease | 2003 | Shiga, Japan | 52 (24 CN, 28 AD)<br>AD grouped by slowly and rapidly declining at follow-up | NINCDS-ADRD A and DSM-III-R | <sup>99m</sup>Tc-HMPAO SPECT | CN: 69.6 (7.2)<br>slowly declining AD: 71.9 (1.0)<br>rapidly declining AD: 71.6 (1.8) | no | no | no | yes |
| Obara et al. | Cognitive declines correlate with decreased cortical volume and perfusion in dementia of Alzheimer type | 1994 | Texas, USA | 36 (18 CN, 18 AD) | NINCDS-ADRD A and DSM-III-R | <sup>133</sup>Xenon CT | CN: 73.7 (7.0)<br>AD: 75.4 (5.2) | yes | yes | no | yes |
| Sase et al. | Discrimination Between Patients With Alzheimer Disease and Healthy Subjects Using | 2017 | Osaka, Japan | 42 (15 CN, 27 AD) | DSM IV | <sup>133</sup>Xe CT | CN: 78.6 (4)<br>AD: 81.7 (3.3) | no | yes | no | yes |
|  | Layer Analysis of Cerebral Blood Flow and Xenon Solubility Coefficient in Xenon-Enhanced Computed Tomography |  |  |  |  |  |  |  |  |  |  |
| Schuff et al. | Cerebral blood flow in ischemic vascular dementia and Alzheimer's disease, measured by arterial spin-labeling magnetic resonance imaging | 2009 | California, USA | 32 (18 CN, 14 AD)<br>also a group with subcortical ischemic vascular dementia | NINCD S-ADRD A and DSM IV | ASL MRI | CN: 73 (8)<br>AD: 74 (5) | yes | yes | no | no |
| Shimizu et al. | Differentiation of dementia with Lewy bodies from Alzheimer's disease using brain SPECT | 2005 | Tokyo, Japan | 103 (28 CN, 75 AD)<br>also a group with dementia with Lewy bodies | NINCD S-ADRD A | <sup>123</sup>I-IMP SPECT | CN: 75.1 (6.4)<br>AD: 75.5 (6.8) | yes | yes | no | no |
| Sundström et al. | Memory-provoked rCBF-SPECT as a diagnostic tool in Alzheimer's disease? | 2006 | Umeå, Sweden | 36 (18 CN, 18 AD) | NINCD S-ADRD A | <sup>99m</sup>Tc-HMPAO SPECT | CN: 69.4 (3.9)<br>AD: 73.2 (4.8)<br>CN are younger | no | yes | no | no |
| Takahashi et al. | Poor performance in Clock-Drawing Test associated with visual memory deficit and reduced bilateral | 2008 | Sagamihara, Japan | 25 (2 CN, 7 MCI, 16 AD)<br>grouped into 11 with high scores and 14 with low scores on the | Petersen et al criteria for MCI; NINDS-ADRD A for AD | <sup>99m</sup>Tc-ECD SPECT | high CDT score: 74.4 (6.8)<br>low CDT score: 75.1 (10) | no | no | no | yes |
|  | hippocampal and left temporoparietal regional blood flows in Alzheimer's disease patients |  |  | Clock Drawing Test (CDT), cutoff 9 out of 10 |  |  |  |  |  |  |  |
| Tateno et al. | Usefulness of a blood flow analyzing program 3DSRT to detect occipital hypoperfusion in dementia with Lewy bodies | 2008 | Sunagawa, Japan | 54 (16 CN, 38 AD)<br>also a group with dementia with Lewy bodies | not clear; the “latest diagnostic criteria” | <sup>99m</sup>Tc-ECD SPECT | CN: 75.8 (5.6)<br>AD: 79.0 (5.3) | yes | yes | no | no |
| van de Haar et al. | Neurovascular unit impairment in early Alzheimer's disease measured with magnetic resonance imaging | 2016 | the Netherlands | 30 (16 CN, 14 MCI/AD) | Dubois criteria for MCI; NINCDS-ADRD A for AD | pCAS L MRI | CN: 76.4, range 65-85<br>MCI/AD: 75.3, range 65-85 | yes | yes | no | no |
| Yew & Nation | Cerebrovascular resistance: effects on cognitive decline, cortical atrophy, and progression to dementia | 2017 | California, USA; ADNI | 232 (112 amyloid-negative CN/MCI, 87 amyloid-positive CN/MCI, 33 AD) | NINCDS-ADRD A for AD; florbeta pir PET with cutoff of SUVR 1.11 for amyloid positivity | PASL MRI | CN/MCI amyloid-negative: 69.2 (0.6)<br>CN/MCI amyloid-positive: 73.7 (0.7)<br>AD: 73.2 (1.2)<br>CN/MCI amyloid-negative group is younger | yes | yes | no | no |
| Yoshida et al. | Protein synthesis in the posterior cingulate cortex in | 2011 | Suita, Japan | 16 (8 CN, 8 AD) | NINCDS-ADRD A | <sup>123</sup>I-IMP SPECT | CN: 72.5 (5.8)<br>AD: 73.0 (5.4) | yes | yes | no | no |
|  | Alzheimer's disease |  |  |  |  |  |  |  |  |  |  |

<sup>123</sup>I-IMP: N-isopropyl-(iodine-123) p-iodoamphetamine, <sup>133</sup>Xe: xenon-133, <sup>99m</sup>Tc: technetium-99m, AD: Alzheimer's disease, *APOE*: apolipoprotein E, ASL: arterial spin labeled, CAMDEX: Cambridge Mental Disorders of the Elderly Examination, CASL: continuous ASL, CBF: cerebral blood flow, CDR: Clinical Dementia Rating scale, CDT: clock drawing test, CN: cognitively normal, CT: computed tomography, DSM: Diagnostic and Statistical Manual of Mental Disorders, ECD: ethyl cysteinate dimer, HMPAO: hexamethylpropyleneamine oxime, ICD-10: International Classification of Diseases Tenth Revision, MCI: mild cognitive impairment, MMSE: Mini-Mental State Exam, MRI: magnetic resonance imaging, NINCDS-ADRD: National Institute of Neurological and Communicative Disorders and Stroke and the Alzheimer's Disease and Related Disorders Association, pCASL: pseudo-continuous ASL, PET: positron emission tomography, SD: standard deviation, SPECT: single photon emission computerized tomography, SPET: single photon emission tomography, SUVR: standardized uptake value ratio

**Supplementary Table 2.**
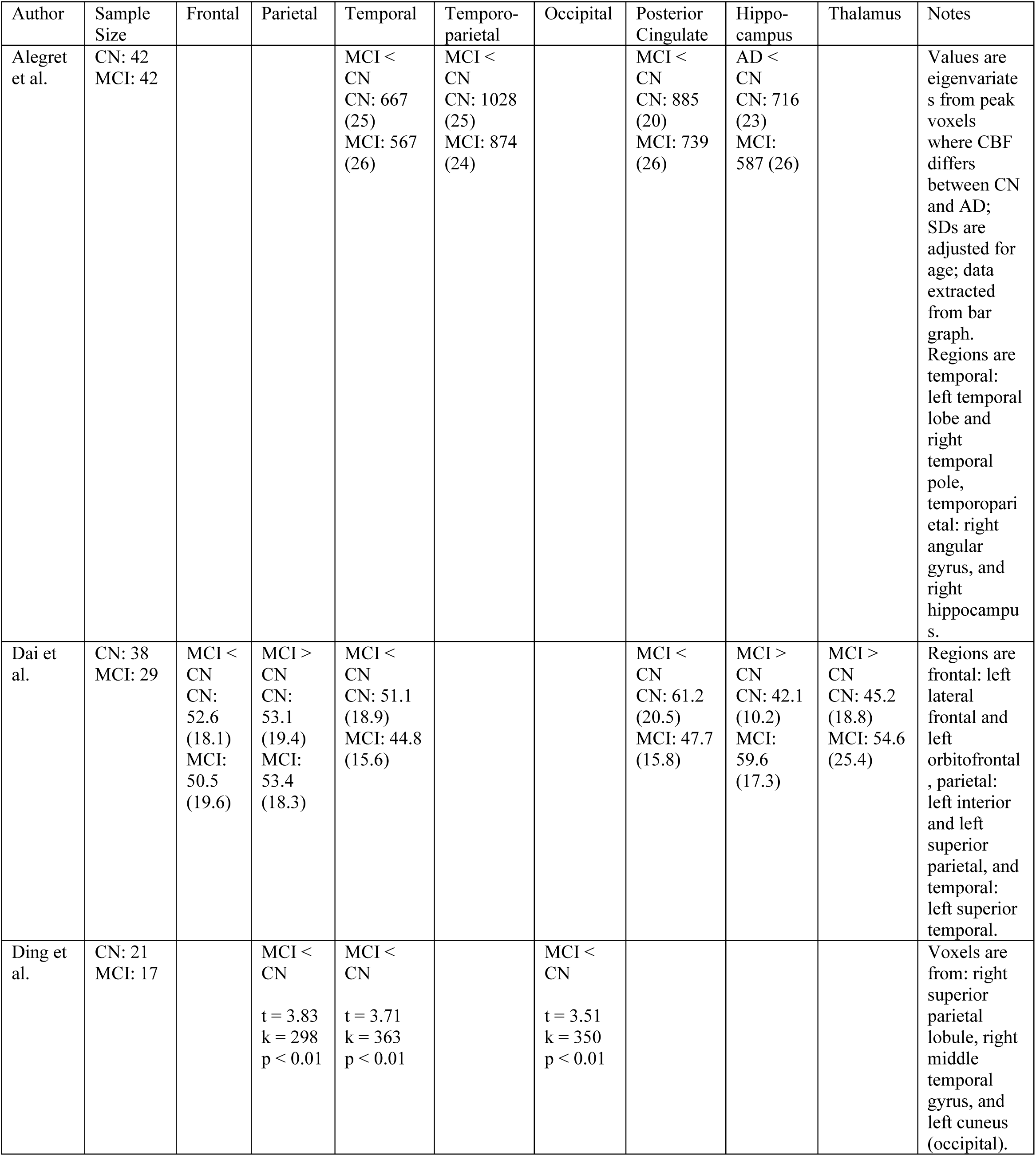
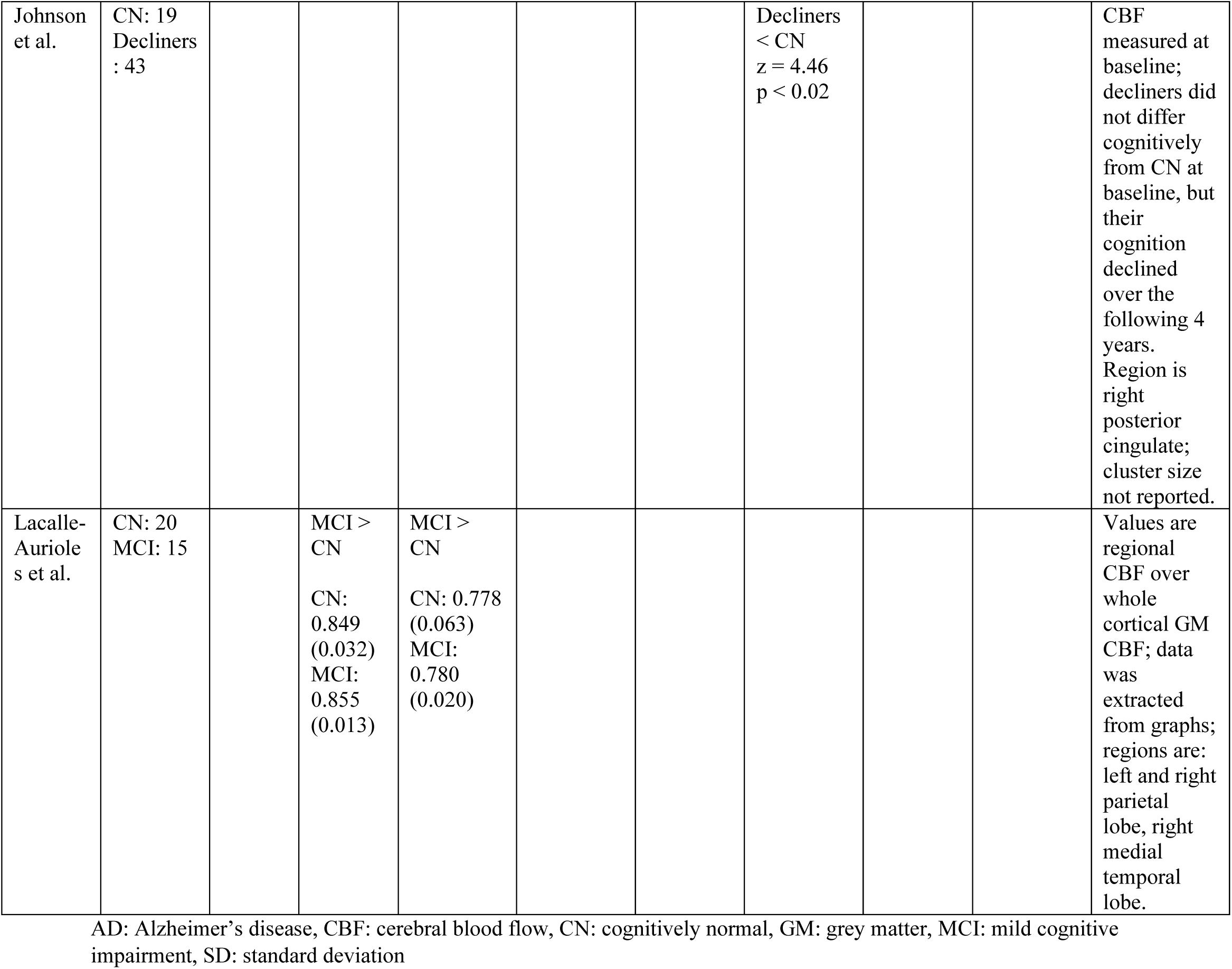
CBF in CN and MCI in each paper.

**Supplementary Table 3.** Relationships between CBF and cognition in each paper.

| Authors | Sample Size | Correlation between CBF and Cognition | Notes |
| --- | --- | --- | --- |
| Alegret et al. | 126 (CN, MCI, & AD) | CBF in posterior cingulate ($r = 0.72$, $p < 0.001$) and right temporal pole ($r = 0.68$, $p < 0.001$) is positively correlated with MMSE score. CBF in posterior cingulate ($r = 0.55$, $p < 0.001$) and right temporal pole ($r = 0.48$, $p < 0.001$) is positively correlated with 15-Objects Test score. CBF in posterior cingulate ($r = 0.46$, $p < 0.001$) and right temporal pole ($r = 0.40$, $p < 0.001$) is positively correlated with 15 item Boston Naming Test score. CBF in posterior cingulate ($r = 0.37$, $p < 0.001$) and right temporal pole ($r = 0.34$, $p < 0.001$) is positively correlated with Poppelreuter test score. | Relationship assessed across all participants; CBF measures are eigenvariates from the named regions. |
| Benoit et al. | 30 AD (with and without apathy) | CBF in bilateral lateral parietal cortex is positively correlated with MMSE score. Significant clusters right: ($z = 3.73$, $p = 0.015$), left: ($z = 3.64$, $p = 0.010$). | Analysis was voxel-wise. Using the apathy score on the Neuropsychiatric Inventory as a covariate in the correlation did not change the results. |
| Brown et al. | 24 AD | Longitudinal change in CAMCOG score is positively correlated with longitudinal change in CBF in right and left low frontal ($r = 0.69$; $r = 0.74$) and right temporal regions ($r = 0.62$). All $p < 0.05$; left low frontal $p < 0.05$ after Bonferroni correction. | Frontal and temporal CBF was positively correlated with language, praxis, and abstraction, but not memory CAMCOG sub-scores in this paper. |
| Firbank et al. | 46 (CN and AD) | Global GM/WM CBF does not contribute to the variation in CAMCOG-R score when age, sex, and hippocampal volume are predictors in the model. |  |
| Jagust et al. | 9 AD | The ratio of CBF in the temporoparietal region: whole tomographic slice is positively correlated with MMSQ score ($r = 0.81$, $p < 0.01$). | |
| Johnson et al. | 124 CN/ MCI | CBF in rostral anterior cingulate and inferior frontal regions was positively correlated with scores on Trailmaking Test Part B ($p < 0.05$), while CBF in caudal anterior cingulate was negatively correlated with scores on the same test ($p < 0.006$). No correlations were seen between anterior cingulate CBF and the California Verbal Learning Test or Self-Ordering Test, or between posterior cingulate CBF and any of the three tests. | |
| Nagahama et al. | 28 AD, both rapidly and slowly progressing | CBF in right middle ($z = 4.67$, $k = 458$, $p < 0.001$) and superior frontal gyrus ($z = 3.05$, $k = 8$, $p < 0.01$) and right inferior parietal cortex ($z = 3.27$, $k = 332$, $p < 0.001$) is decreased in patients that decline rapidly compared to those that decline slowly, meaning that their MMSE score declines less than 4 points over 2 to 3 years of follow-up. | Results reported from voxel-wise analysis. |
| Obara et al. | 17 AD | Declines in parietal cortical CBF add to the prediction of cognitive decline on the CCSE ($p = 0.015$) when other predictors are decline in cortical volumes and decline in subcortical densities. | CBF, cognition, and tissue volumes and densities were measured every 6 to 12 months for between 2 and 3 years on average. |
| Sase et al. | 42 (CN and AD) | CBF: lambda ratio in the third ($r = 0.66$, $p < 0.0001$) and fourth ($r = 0.68$, $p < 0.0001$) layer left lateral views is positively correlated with MMSE scores. | Lambda is the Xenon solubility coefficient. CBF and CBF:lambda ratio were decreased in AD compared to CN in this paper. |
| Takahashi et al. | 25 (CN, MCI, and AD) | Clock Drawing Test scores are positively correlated with CBF in the left hippocampus ($\rho = 0.43$, $p < 0.05$), left parietal ($\rho = 0.37$), bilateral pericallosal ($\rho = 0.35$; $0.32$), and bilateral angular regions ($\rho = 0.38$; $0.35$). | Correlation coefficients are Spearman's rho. Those without p values did not reach significance. Areas that with correlations less than $\rho = 0.32$ were: callosomarginal, precentral, central, right parietal, temporal, posterior, lenticular nucleus, thalamus, right hippocampus, and cerebellum. |

AD: Alzheimer's disease, CAMCOG: Cambridge Cognitive Examination, CAMCOG-R: CAMCOG Revised, CBF: cerebral blood flow, CCSE: Cognitive Capacity Screening Examination, CN: cognitively normal, GM: grey matter, MCI: mild cognitive impairment, MMSE: Mini-Mental State Exam, MMSQ: Mini-Mental State Questionnaire, WM: white matter

## References

1. 2021 Alzheimer’s disease facts and figures. (2021). Alzheimers Dement, 17(3), 327–406. https://doi.org/10.1002/alz.12328

2. Alegret, M., Vinyes-Junqué, G., Boada, M., Martínez-Lage, P., Cuberas, G., Espinosa, A., Roca, I., Hernández, I., Valero, S., Rosende-Roca, M., Mauleón, A., Becker, Jt, & Tárraga, L. (2010). Brain perfusion correlates of visuoperceptual deficits in mild cognitive impairment and mild Alzheimer’s disease. J Alzheimers Dis, 21(2), 557–567. https://doi.org/10.3233/jad-2010-091069

3. Benoit, M., Koulibaly, P. M., Migneco, O., Darcourt, J., Pringuey, D. J., & Robert, P. H. (2002). Brain perfusion in Alzheimer’s disease with and without apathy: a SPECT study with statistical parametric mapping analysis. Psychiatry Res, 114(2), 103–111. https://doi.org/10.1016/s0925-4927(02)00003-3

4. Blennow, K., & Zetterberg, H. (2018). The Past and the Future of Alzheimer’s Disease Fluid Biomarkers. J Alzheimers Dis, 62(3), 1125–1140. https://doi.org/10.3233/JAD-170773

5. Brown, D. R., Hunter, R., Wyper, D. J., Patterson, J., Kelly, R. C., Montaldi, D., & McCullouch, J. (1996). Longitudinal changes in cognitive function and regional cerebral function in Alzheimer’s disease: a SPECT blood flow study. J Psychiatr Res, 30(2), 109–126. https://doi.org/10.1016/0022-3956(95)00032-1

6. Brown, D. R., Wyper, D. J., Owens, J., Patterson, J., Kelly, R. C., Hunter, R., & McCulloch, J. (1997). 123Iodo-MK-801: a spect agent for imaging the pattern and extent of glutamate (NMDA) receptor activation in Alzheimer’s disease. J Psychiatr Res, 31(6), 605–619. https://doi.org/10.1016/s0022-3956(97)00031-9

7. Cho, H., Kwon, J. H., Seo, H. J., & Kim, J. S. (2010). The short-term effect of acetylcholinesterase inhibitor on the regional cerebral blood flow of Alzheimer’s disease. Arch Gerontol Geriatr, 50(2), 222–226. https://doi.org/10.1016/j.archger.2009.03.013

8. Claus, J. J., van Harskamp, F., Breteler, M. M., Krenning, E. P., van der Cammen, T. J., Hofman, A., & Hasan, D. (1994). Assessment of cerebral perfusion with single-photon emission tomography in normal subjects and in patients with Alzheimer’s disease: effects of region of interest selection. Eur J Nucl Med, 21(10), 1044–1051. https://doi.org/10.1007/bf00181058

9. Colloby, S. J., Fenwick, J. D., Williams, E. D., Paling, S. M., Lobotesis, K., Ballard, C., McKeith, I., & O’Brien, J. T. (2002). A comparison of (99m)Tc-HMPAO SPET changes in dementia with Lewy bodies and Alzheimer’s disease using statistical parametric mapping. Eur J Nucl Med Mol Imaging, 29(5), 615–622. https://doi.org/10.1007/s00259-002-0778-5

10. Counts, S. E., Ikonomovic, M. D., Mercado, N., Vega, I. E., & Mufson, E. J. (2017). Biomarkers for the Early Detection and Progression of Alzheimer’s Disease. Neurotherapeutics, 14(1), 35–53. https://doi.org/10.1007/s13311-016-0481-z

11. Culpepper, L., Lam, R. W., & McIntyre, R. S. (2017). Cognitive Impairment in Patients With Depression: Awareness, Assessment, and Management. J Clin Psychiatry, 78(9), 1383–1394. https://doi.org/10.4088/JCP.tk16043ah5c

12. Dai, W., Lopez, O. L., Carmichael, O. T., Becker, J. T., Kuller, L. H., & Gach, H. M. (2009). Mild cognitive impairment and alzheimer disease: patterns of altered cerebral blood flow at MR imaging. Radiology, 250(3), 856–866. https://doi.org/10.1148/radiol.2503080751

13. De Reuck, J., Decoo, D., Van Aken, J., Strijckmans, K., Lemahieu, I., & Vermeulen, A. (1992). Positron emission tomography study of the human hypothalamus during normal ageing and in ischemic and degenerative disorders. Clin Neurol Neurosurg, 94(2), 113–118. https://doi.org/10.1016/0303-8467(92)90067-d

14. Detre, J. A., Rao, H., Wang, D. J., Chen, Y. F., & Wang, Z. (2012). Applications of arterial spin labeled MRI in the brain. J Magn Reson Imaging, 35(5), 1026–1037. https://doi.org/10.1002/jmri.23581

15. Ding, B., Ling, H. W., Zhang, Y., Huang, J., Zhang, H., Wang, T., & Yan, F. H. (2014). Pattern of cerebral hyperperfusion in Alzheimer’s disease and amnestic mild cognitive impairment using voxel-based analysis of 3D arterial spin-labeling imaging: initial experience. Clin Interv Aging, 9, 493–500. https://doi.org/10.2147/cia.S58879

16. Dougall, N., Nobili, F., & Ebmeier, K. P. (2004). Predicting the accuracy of a diagnosis of Alzheimer’s disease with 99mTc HMPAO single photon emission computed tomography. Psychiatry Res, 131(2), 157–168. https://doi.org/10.1016/j.pscychresns.2003.11.001

17. Duan, W., Zhou, G. D., Balachandrasekaran, A., Bhumkar, A. B., Boraste, P. B., Becker, J. T., Kuller, L. H., Lopez, O. L., Gach, H. M., & Dai, W. (2021). Cerebral Blood Flow Predicts Conversion of Mild Cognitive Impairment into Alzheimer’s Disease and Cognitive Decline: An Arterial Spin Labeling Follow-up Study. J Alzheimers Dis, 82(1), 293–305. https://doi.org/10.3233/JAD-210199

18. Edelman, R. R., Mattle, H. P., Atkinson, D. J., Hill, T., Finn, J. P., Mayman, C., Ronthal, M., Hoogewoud, H. M., & Kleefield, J. (1990). Cerebral blood flow: assessment with dynamic contrast-enhanced T2*-weighted MR imaging at 1.5 T. Radiology, 176(1), 211–220. https://doi.org/10.1148/radiology.176.1.2353094

19. Encinas, M., De Juan, R., Marcos, A., Gil, P., Barabash, A., Fernández, C., De Ugarte, C., & Cabranes, J. A. (2003). Regional cerebral blood flow assessed with 99mTc-ECD SPET as a marker of progression of mild cognitive impairment to Alzheimer’s disease. Eur J Nucl Med Mol Imaging, 30(11), 1473–1480. https://doi.org/10.1007/s00259-003-1277-z

20. Firbank, M. J., He, J., Blamire, A. M., Singh, B., Danson, P., Kalaria, R. N., & O’Brien, J. T. (2011). Cerebral blood flow by arterial spin labeling in poststroke dementia. Neurology, 76(17), 1478–1484. https://doi.org/10.1212/WNL.0b013e318217e76a

21. Haji, M., Kimura, N., Hanaoka, T., Aso, Y., Takemaru, M., Hirano, T., & Matsubara, E. (2015). Evaluation of regional cerebral blood flow in Alzheimer’s disease patients with subclinical hypothyroidism. Dement Geriatr Cogn Disord, 39(5-6), 360–367. https://doi.org/10.1159/000375298

22. Hanyu, H., Shimuzu, T., Tanaka, Y., Takasaki, M., Koizumi, K., & Abe, K. (2003). Effect of age on regional cerebral blood flow patterns in Alzheimer’s disease patients. J Neurol Sci, 209(1-2), 25–30. https://doi.org/10.1016/s0022-510x(02)00456-2

23. Hanyu, H., Shimizu, S., Tanaka, Y., Takasaki, M., Koizumi, K., & Abe, K. (2004). Differences in regional cerebral blood flow patterns in male versus female patients with Alzheimer disease. AJNR Am J Neuroradiol, 25(7), 1199–1204.

24. Hanyu, H., Sato, T., Shimizu, S., Kanetaka, H., Iwamoto, T., & Koizumi, K. (2008). The effect of education on rCBF changes in Alzheimer’s disease: a longitudinal SPECT study. Eur J Nucl Med Mol Imaging, 35(12), 2182–2190. https://doi.org/10.1007/s00259-008-0848-4

25. Hardy, J. A., & Higgins, G. A. (1992). Alzheimer’s disease: the amyloid cascade hypothesis. Science, 256(5054), 184–185. https://doi.org/10.1126/science.1566067

26. Harris, G. J., Links, J. M., Pearlson, G. D., & Camargo, E. E. (1991). Cortical circumferential profile of SPECT cerebral perfusion in Alzheimer’s disease. Psychiatry Res, 40(3), 167–180. https://doi.org/10.1016/0925-4927(91)90008-e

27. Iadecola, C., & Gottesman, R. F. (2019). Neurovascular and Cognitive Dysfunction in Hypertension. Circ Res, 124(7), 1025–1044. https://doi.org/10.1161/CIRCRESAHA.118.313260

28. Iizuka, T., & Kameyama, M. (2017). Cholinergic enhancement increases regional cerebral blood flow to the posterior cingulate cortex in mild Alzheimer’s disease. Geriatr Gerontol Int, 17(6), 951–958. https://doi.org/10.1111/ggi.12818

29. Iturria-Medina, Y., Carbonell, F. M., Sotero, R. C., Chouinard-Decorte, F., & Evans, A. C. (2017). Multifactorial causal model of brain (dis)organization and therapeutic intervention: Application to Alzheimer’s disease. NeuroImage, 152, 60–77. https://doi.org/10.1016/j.neuroimage.2017.02.058

30. Jagust, W. J., Budinger, T. F., & Reed, B. R. (1987). The diagnosis of dementia with single photon emission computed tomography. Arch Neurol, 44(3), 258–262. https://doi.org/10.1001/archneur.1987.00520150014011

31. Jagust, W. J., Reed, B. R., Seab, J. P., & Budinger, T. F. (1990). Alzheimer’s disease. Age at onset and single-photon emission computed tomographic patterns of regional cerebral blood flow. Arch Neurol, 47(6), 628–633. https://doi.org/10.1001/archneur.1990.00530060036013

32. Jessen, F., Amariglio, R. E., van Boxtel, M., Breteler, M., Ceccaldi, M., Chetelat, G., Dubois, B., Dufouil, C., Ellis, K. A., van der Flier, W. M., Glodzik, L., van Harten, A. C., de Leon, M. J., McHugh, P., Mielke, M. M., Molinuevo, J. L., Mosconi, L., Osorio, R. S., Perrotin, A., Petersen, R. C., Rabin, L. A., Rami, L., Reisberg, B., Rentz, D. M., Sachdev, P. S., de la Sayette, V., Saykin, A. J., Scheltens, P., Shulman, M. B., Slavin, M. J., Sperling, R. A., Stewart, R., Uspenskaya, O., Vellas, B., Visser, P. J., Wagner, M., & Subjective Cognitive Decline Initiative Working, G. (2014). A conceptual framework for research on subjective cognitive decline in preclinical Alzheimer’s disease. Alzheimers Dement, 10(6), 844–852. https://doi.org/10.1016/j.jalz.2014.01.001

33. Jessen, F., Amariglio, R. E., Buckley, R. F., van der Flier, W. M., Han, Y., Molinuevo, J. L., Rabin, L., Rentz, D. M., Rodriguez-Gomez, O., Saykin, A. J., Sikkes, S. A. M., Smart, C. M., Wolfsgruber, S., & Wagner, M. (2020). The characterisation of subjective cognitive decline. Lancet Neurol, 19(3), 271–278. https://doi.org/10.1016/S1474-4422(19)30368-0

34. Johnson, K. A., Moran, E. K., Becker, J. A., Blacker, D., Fischman, A. J., & Albert, M. S. (2007). Single photon emission computed tomography perfusion differences in mild cognitive impairment. J Neurol Neurosurg Psychiatry, 78(3), 240–247. https://doi.org/10.1136/jnnp.2006.096800

35. Jones, T., Chesler, D. A., & Ter-Pogossian, M. M. (1976). The continuous inhalation of oxygen-15 for assessing regional oxygen extraction in the brain of man. Br J Radiol, 49(580), 339–343. https://doi.org/10.1259/0007-1285-49-580-339

36. Kawamura, J., Meyer, J. S., Terayama, Y., & Weathers, S. (1991). Cerebral white matter perfusion in dementia of Alzheimer type. Alzheimer Dis Assoc Disord, 5(4), 231–239. https://doi.org/10.1097/00002093-199100540-00002

37. Kety, S. S., & Schmidt, C. F. (1948). The Nitrous Oxide Method for the Quantitative Determination of Cerebral Blood Flow in Man: Theory, Procedure and Normal Values. J Clin Invest, 27(4), 476–483. https://doi.org/10.1172/JCI101994

38. Kimura, N., Kumamoto, T., Masuda, H., Hanaoka, T., Hazama, Y., Okazaki, T., & Arakawa, R. (2011). Relationship between thyroid hormone levels and regional cerebral blood flow in Alzheimer disease. Alzheimer Dis Assoc Disord, 25(2), 138–143. https://doi.org/10.1097/WAD.0b013e3181f9aff2

39. Lacalle-Aurioles, M., Alemán-Gómez, Y., Guzmán-De-Villoria, J. A., Cruz-Orduña, I., Olazarán, J., Mateos-Pérez, J. M., Martino, M. E., & Desco, M. (2013). Is the cerebellum the optimal reference region for intensity normalization of perfusion MR studies in early Alzheimer’s disease? PLoS One, 8(12), e81548. https://doi.org/10.1371/journal.pone.0081548

40. Lassen, N. A., & Ingvar, D. H. (1961). The blood flow of the cerebral cortex determined by radioactive krypton. Experientia, 17, 42–43. https://doi.org/10.1007/BF02157946

41. Lassen, N. A., Hoedt-Rasmussen, K., Sorensen, S. C., Skinhoj, E., Cronquist, S., Bodforss, B., & Ingvar, D. H. (1963). Regional Cerebral Blood Flow in Man Determined by Krypton. Neurology, 13, 719–727. https://doi.org/10.1212/wnl.13.9.719

42. Lee, Y. C., Liu, R. S., Liao, Y. C., Sun, C. M., Wang, P. S., Wang, P. N., & Liu, H. C. (2003). Statistical parametric mapping of brain SPECT perfusion abnormalities in patients with Alzheimer’s disease. Eur Neurol, 49(3), 142–145. https://doi.org/10.1159/000069086

43. Maeda, M., Itoh, S., Kimura, H., Iwasaki, T., Hayashi, N., Yamamoto, K., Ishii, Y., & Kubota, T. (1993). Tumor vascularity in the brain: evaluation with dynamic susceptibility-contrast MR imaging. Radiology, 189(1), 233–238. https://doi.org/10.1148/radiology.189.1.8372199

44. Matsuda, H. (2001). Cerebral blood flow and metabolic abnormalities in Alzheimer’s disease. Ann Nucl Med, 15(2), 85–92. https://doi.org/10.1007/BF02988596

45. Moretti, D. V. (2016). Electroencephalography-driven approach to prodromal Alzheimer’s disease diagnosis: from biomarker integration to network-level comprehension. Clin Interv Aging, 11, 897–912. https://doi.org/10.2147/cia.S103313

46. Mortel, K. F., Pavol, M. A., Wood, S., Meyer, J. S., Terayama, Y., Rexer, J. L., & Herod, B. (1994). Prospective studies of cerebral perfusion and cognitive testing among elderly normal volunteers and patients with ischemic vascular dementia and Alzheimer’s disease. Angiology, 45(3), 171–180. https://doi.org/10.1177/000331979404500301

47. Mubrin, Z., Knezevic, S., Spilich, G., Risberg, J., Gubarev, N., Wannenmacher, W., & Vucinic, G. (1989). Normalization of rCBF pattern in senile dementia of the Alzheimer’s type. Psychiatry Res, 29(3), 303–306. https://doi.org/10.1016/0165-1781(89)90072-3

48. Nagahama, Y., Nabatame, H., Okina, T., Yamauchi, H., Narita, M., Fujimoto, N., Murakami, M., Fukuyama, H., & Matsuda, M. (2003). Cerebral correlates of the progression rate of the cognitive decline in probable Alzheimer’s disease. Eur Neurol, 50(1), 1–9. https://doi.org/10.1159/000070851

49. Obara, K., Meyer, J. S., Mortel, K. F., & Muramatsu, K. (1994). Cognitive declines correlate with decreased cortical volume and perfusion in dementia of Alzheimer type. J Neurol Sci, 127(1), 96–102. https://doi.org/10.1016/0022-510x(94)90141-4

50. Pearlson, G. D., Harris, G. J., Powers, R. E., Barta, P. E., Camargo, E. E., Chase, G. A., Noga, J. T., & Tune, L. E. (1992). Quantitative changes in mesial temporal volume, regional cerebral blood flow, and cognition in Alzheimer’s disease. Arch Gen Psychiatry, 49(5), 402–408. https://doi.org/10.1001/archpsyc.1992.01820050066012

51. Petersen, R. C., Smith, G. E., Waring, S. C., Ivnik, R. J., Tangalos, E. G., & Kokmen, E. (1999). Mild cognitive impairment: clinical characterization and outcome. Arch Neurol, 56(3), 303–308. https://doi.org/10.1001/archneur.56.3.303

52. Petersen, R. C. (2016). Mild Cognitive Impairment. Continuum (Minneap Minn), 22(2 Dementia), 404–418. https://doi.org/10.1212/CON.0000000000000313

53. Rempp, K. A., Brix, G., Wenz, F., Becker, C. R., Guckel, F., & Lorenz, W. J. (1994). Quantification of regional cerebral blood flow and volume with dynamic susceptibility contrast-enhanced MR imaging. Radiology, 193(3), 637–641. https://doi.org/10.1148/radiology.193.3.7972800

54. Richardson, W. S., Wilson, M. C., Nishikawa, J., & Hayward, R. S. (1995). The well-built clinical question: a key to evidence-based decisions. ACP J Club, 123(3), A12–13. https://www.ncbi.nlm.nih.gov/pubmed/7582737

55. Sase, S., Yamamoto, H., Kawashima, E., Tan, X., & Sawa, Y. (2017). Discrimination Between Patients With Alzheimer Disease and Healthy Subjects Using Layer Analysis of Cerebral Blood Flow and Xenon Solubility Coefficient in Xenon-Enhanced Computed Tomography. J Comput Assist Tomogr, 41(3), 477–483. https://doi.org/10.1097/rct.0000000000000525

56. Schuff, N., Matsumoto, S., Kmiecik, J., Studholme, C., Du, A., Ezekiel, F., Miller, B. L., Kramer, J. H., Jagust, W. J., Chui, H. C., & Weiner, M. W. (2009). Cerebral blood flow in ischemic vascular dementia and Alzheimer’s disease, measured by arterial spin-labeling magnetic resonance imaging. Alzheimers Dement, 5(6), 454–462. https://doi.org/10.1016/j.jalz.2009.04.1233

57. Shimizu, S., Hanyu, H., Kanetaka, H., Iwamoto, T., Koizumi, K., & Abe, K. (2005). Differentiation of dementia with Lewy bodies from Alzheimer’s disease using brain SPECT. Dement Geriatr Cogn Disord, 20(1), 25–30. https://doi.org/10.1159/000085070

58. Starkstein, S. E., Migliorelli, R., Tesón, A., Sabe, L., Vázquez, S., Turjanski, M., Robinson, R. G., & Leiguarda, R. (1994). Specificity of changes in cerebral blood flow in patients with frontal lobe dementia. J Neurol Neurosurg Psychiatry, 57(7), 790–796. https://doi.org/10.1136/jnnp.57.7.790

59. Sundström, T., Elgh, E., Larsson, A., Näsman, B., Nyberg, L., & Riklund, K. A. (2006). Memory-provoked rCBF-SPECT as a diagnostic tool in Alzheimer’s disease? Eur J Nucl Med Mol Imaging, 33(1), 73–80. https://doi.org/10.1007/s00259-005-1874-0

60. Takahashi, M., Sato, A., Nakajima, K., Inoue, A., Oishi, S., Ishii, T., & Miyaoka, H. (2008). Poor performance in Clock-Drawing Test associated with visual memory deficit and reduced bilateral hippocampal and left temporoparietal regional blood flows in Alzheimer’s disease patients. Psychiatry Clin Neurosci, 62(2), 167–173. https://doi.org/10.1111/j.1440-1819.2008.01750.x

61. Tateno, M., Kobayashi, S., Shirasaka, T., Furukawa, Y., Fujii, K., Morii, H., Yasumura, S., Utsumi, K., & Saito, T. (2008). Comparison of the usefulness of brain perfusion SPECT and MIBG myocardial scintigraphy for the diagnosis of dementia with Lewy bodies. Dement Geriatr Cogn Disord, 26(5), 453–457. https://doi.org/10.1159/000165918

62. van de Haar, H. J., Jansen, J. F. A., van Osch, M. J. P., van Buchem, M. A., Muller, M., Wong, S. M., Hofman, P. A. M., Burgmans, S., Verhey, F. R. J., & Backes, W. H. (2016). Neurovascular unit impairment in early Alzheimer’s disease measured with magnetic resonance imaging. Neurobiol Aging, 45, 190–196. https://doi.org/10.1016/j.neurobiolaging.2016.06.006

63. Vavilala, M. S., Lee, L. A., & Lam, A. M. (2002). Cerebral blood flow and vascular physiology. Anesthesiol Clin North Am, 20(2), 247–264, v. https://doi.org/10.1016/s0889-8537(01)00012-8

64. Vogel, A., Hasselbalch, S. G., Gade, A., Ziebell, M., & Waldemar, G. (2005). Cognitive and functional neuroimaging correlate for anosognosia in mild cognitive impairment and Alzheimer’s disease. Int J Geriatr Psychiatry, 20(3), 238–246. https://doi.org/10.1002/gps.1272

65. Warwick, J. M. (2004). Imaging of brain function using SPECT. Metab Brain Dis, 19(1-2), 113–123. https://doi.org/10.1023/b:mebr.0000027422.48744.a3

66. Yew, B., & Nation, D. A. (2017). Cerebrovascular resistance: effects on cognitive decline, cortical atrophy, and progression to dementia. Brain, 140(7), 1987–2001. https://doi.org/10.1093/brain/awx112

67. Yoshida, T., Kazui, H., Tokunaga, H., Kito, Y., Kubo, Y., Kimura, N., Morihara, T., Shimosegawa, E., Hatazawa, J., & Takeda, M. (2011). Protein synthesis in the posterior cingulate cortex in Alzheimer’s disease. Psychogeriatrics, 11(1), 40–45. https://doi.org/10.1111/j.1479-8301.2010.00350.x

68. Zhang, H., Wang, Y., Lyu, D., Li, Y., Li, W., Wang, Q., Li, Y., Qin, Q., Wang, X., Gong, M., Jiao, H., Liu, W., & Jia, J. (2021). Cerebral blood flow in mild cognitive impairment and Alzheimer’s disease: A systematic review and meta-analysis. Ageing Res Rev, 71, 101450. https://doi.org/10.1016/j.arr.2021.101450

69. Zhu, C. C., Fu, S. Y., Chen, Y. X., Li, L., Mao, R. L., Wang, J. Z., Liu, R., Liu, Y., & Wang, X. C. (2020). Advances in Drug Therapy for Alzheimer’s Disease. Curr Med Sci, 40(6), 999–1008. https://doi.org/10.1007/s11596-020-2281-2

70. Zlokovic, B. V. (2011). Neurovascular pathways to neurodegeneration in Alzheimer’s disease and other disorders. Nat Rev Neurosci, 12(12), 723–738. https://doi.org/10.1038/nrn3114

