## Supplementary material for "Altered Cerebral Blood Flow in Older Adults with Alzheimer’s Disease: A Systematic Review": PRISMA Checklist

### PRISMA 2020 expanded checklist

Note: This expanded checklist details elements recommended for reporting for each PRISMA 2020 item. Non-italicized elements are considered 'essential' and should be reported in the main report or as supplementary material for all systematic reviews (except for those preceded by "If…", which should only be reported where applicable). Elements written in italics are 'additional', and while not essential, provide supplementary information that may enhance the completeness and usability of systematic review reports. Note that elements presented here are an abridged version of those presented in the explanation and elaboration paper (BMJ 2021;372:n160), with references and some examples removed. Consulting the explanation and elaboration paper is recommended if further clarity or information is required.

| Section and Topic | Item # | Elements recommended for reporting |
|---|---|---|
| **TITLE** | | |
| TITLE | 1 | • Identify the report as a systematic review in the title.<br>• Report an informative title that provides key information about the main objective or question the review addresses (e.g. the population(s) and intervention(s) the review addresses).<br>• *Consider providing additional information in the title, such as the method of analysis used, the designs of included studies, or an indication that the review is an update of an existing review, or a continually updated ("living") systematic review.* |
| **ABSTRACT** | | |
| ABSTRACT | 2 | • Report an abstract addressing each item in the PRISMA 2020 for Abstracts checklist. |
| **INTRODUCTION** | | |
| RATIONALE | 3 | • Describe the current state of knowledge and its uncertainties.<br>• Articulate why it is important to do the review.<br>• If other systematic reviews addressing the same (or a largely similar) question are available, explain why the current review was considered necessary. If the review is an update or replication of a particular systematic review, indicate this and cite the previous review.<br>• If the review examines the effects of interventions, also briefly describe how the intervention(s) examined might work.<br>• *If there is complexity in the intervention or context of its delivery (or both) (e.g. multi-component interventions, equity considerations), consider presenting a logic model to visually display the hypothesised relationship between intervention components and outcomes.* |
| OBJECTIVES | 4 | • Provide an explicit statement of all objective(s) or question(s) the review addresses, expressed in terms of a relevant question formulation framework.<br>• If the purpose is to evaluate the effects of interventions, use the Population, Intervention, Comparator, Outcome (PICO) framework or one of its variants, to state the comparisons that will be made. |
| **METHODS** | | |
| ELIGIBILITY CRITERIA | 5 | • Specify all study characteristics used to decide whether a study was eligible for inclusion in the review, that is, components described in the PICO framework or one of its variants, and other characteristics, such as eligible study design(s) and setting(s), and minimum duration of follow-up.<br>• Specify eligibility criteria with regard to report characteristics, such as year of dissemination, language, and report status (e.g. whether reports, such as unpublished manuscripts and conference abstracts, were eligible for inclusion).<br>• Clearly indicate if studies were ineligible because the outcomes of interest were not measured, or ineligible because the results for the outcome of interest were not reported.<br>• Specify any groups used in the synthesis (e.g. intervention, outcome and population groups) and link these to the comparisons specified in the objectives (item #4).<br>• *Consider providing rationales for any notable restrictions to study eligibility.* |

| Section and Topic | Item # | Elements recommended for reporting |
|---|---|---|
| INFORMATION SOURCES | 6 | • Specify the date when each source (e.g. database, register, website, organisation) was last searched or consulted.<br>• If bibliographic databases were searched, specify for each database its name (e.g. MEDLINE, CINAHL), the interface or platform through which the database was searched (e.g. Ovid, EBSCOhost), and the dates of coverage (where this information is provided).<br>• If study registers, regulatory databases and other online repositories were searched, specify the name of each source and any date restrictions that were applied.<br>• If websites, search engines or other online sources were browsed or searched, specify the name and URL of each source.<br>• If organisations or manufacturers were contacted to identify studies, specify the name of each source.<br>• If individuals were contacted to identify studies, specify the types of individuals contacted (e.g. authors of studies included in the review or researchers with expertise in the area).<br>• If reference lists were examined, specify the types of references examined (e.g. references cited in study reports included in the systematic review, or references cited in systematic review reports on the same or similar topic).<br>• If cited or citing reference searches (also called backward and forward citation searching) were conducted, specify the bibliographic details of the reports to which citation searching was applied, the citation index or platform used (e.g. Web of Science), and the date the citation searching was done.<br>• If journals or conference proceedings were consulted, specify of the names of each source, the dates covered and how they were searched (e.g. handsearching or browsing online). |
| SEARCH STRATEGY | 7 | • Provide the full line by line search strategy as run in each database with a sophisticated interface (such as Ovid), or the sequence of terms that were used to search simpler interfaces, such as search engines or websites.<br>• Describe any limits applied to the search strategy (e.g. date or language) and justify these by linking back to the review's eligibility criteria.<br>• If published approaches, including search filters designed to retrieve specific types of records or search strategies from other systematic reviews, were used, cite them. If published approaches were adapted, for example if search filters are amended, note the changes made.<br>• If natural language processing or text frequency analysis tools were used to identify or refine keywords, synonyms or subject indexing terms to use in the search strategy, specify the tool(s) used.<br>• If a tool was used to automatically translate search strings for one database to another, specify the tool used.<br>• If the search strategy was validated, for example by evaluating whether it could identify a set of clearly eligible studies, report the validation process used and specify which studies were included in the validation set.<br>• If the search strategy was peer reviewed, report the peer review process used and specify any tool used such as the Peer Review of Electronic Search Strategies (PRESS) checklist.<br>• If the search strategy structure adopted was not based on a PICO-style approach, describe the final conceptual structure and any explorations that were undertaken to achieve it. |
| SELECTION PROCESS | 8 | *Recommendations for reporting regardless of the selection processes used:*<br>• Report how many reviewers screened each record (title/abstract) and each report retrieved, whether multiple reviewers worked independently at each stage of screening or not, and any processes used to resolve disagreements between screeners.<br>• Report any processes used to obtain or confirm relevant information from study investigators.<br>• If abstracts or articles required translation into another language to determine their eligibility, report how these were translated.<br>*Recommendations for reporting in systematic reviews using automation tools in the selection process:*<br>• Report how automation tools were integrated within the overall study selection process. |

| Section and Topic | Item # | Elements recommended for reporting |
|---|---|---|
| | | • If an externally derived machine learning classifier was applied (e.g. Cochrane RCT Classifier), either to eliminate records or to replace a single screener, include a reference or URL to the version used. If the classifier was used to eliminate records *before screening*, report the number eliminated in the PRISMA flow diagram as 'Records marked as ineligible by automation tools'.<br>• If an internally derived machine learning classifier was used to assist with the screening process, identify the software/classifier and version, describe how it was used (e.g. to remove records or replace a single screener) and trained (if relevant), and what internal or external validation was done to understand the risk of missed studies or incorrect classifications.<br>• If machine learning algorithms were used to prioritise screening (whereby unscreened records are continually re-ordered based on screening decisions), state the software used and provide details of any screening rules applied.<br>*Recommendations for reporting in systematic reviews using crowdsourcing or previous 'known' assessments in the selection process:*<br>• If crowdsourcing was used to screen records, provide details of the platform used and specify how it was integrated within the overall study selection process.<br>• If datasets of already-screened records were used to eliminate records retrieved by the search from further consideration, briefly describe the derivation of these datasets. |
| DATA COLLECTION PROCESS | 9 | • Report how many reviewers collected data from each report, whether multiple reviewers worked independently or not, and any processes used to resolve disagreements between data collectors.<br>• Report any processes used to obtain or confirm relevant data from study investigators.<br>• If any automation tools were used to collect data, report how the tool was used, how the tool was trained, and what internal or external validation was done to understand the risk of incorrect extractions.<br>• If articles required translation into another language to enable data collection, report how these articles were translated.<br>• If any software was used to extract data from figures, specify the software used.<br>• If any decision rules were used to select data from multiple reports corresponding to a study, and any steps were taken to resolve inconsistencies across reports, report the rules and steps used. |
| DATA ITEMS (outcomes) | 10a | • List and define the outcome domains and time frame of measurement for which data were sought.<br>• Specify whether all results that were compatible with each outcome domain in each study were sought, and if not, what process was used to select results within eligible domains.<br>• If any changes were made to the inclusion or definition of the outcome domains, or to the importance given to them in the review, specify the changes, along with a rationale.<br>• If any changes were made to the processes used to select results within eligible outcome domains, specify the changes, along with a rationale.<br>• *Consider specifying which outcome domains were considered the most important for interpreting the review's conclusions and provide rationale for the labelling (e.g. "a recent core outcome set identified the outcomes labelled 'critical' as being the most important to patients").* |
| DATA ITEMS (other variables) | 10b | • List and define all other variables for which data were sought (e.g. participant and intervention characteristics, funding sources).<br>• Describe any assumptions made about any missing or unclear information from the studies.<br>• If a tool was used to inform which data items to collect, cite the tool used. |
| STUDY RISK OF BIAS ASSESSMENT | 11 | • Specify the tool(s) (and version) used to assess risk of bias in the included studies.<br>• Specify the methodological domains/components/items of the risk of bias tool(s) used.<br>• Report whether an overall risk of bias judgement that summarised across domains/components/items was made, and if so, what rules were used to reach an overall judgement. |

| Section and Topic | Item # | Elements recommended for reporting |
|---|---|---|
| | | • If any adaptations to an existing tool to assess risk of bias in studies were made, specify the adaptations.<br>• If a new risk of bias tool was developed for use in the review, describe the content of the tool and make it publicly accessible.<br>• Report how many reviewers assessed risk of bias in each study, whether multiple reviewers worked independently, and any processes used to resolve disagreements between assessors.<br>• Report any processes used to obtain or confirm relevant information from study investigators.<br>• If an automation tool was used to assess risk of bias, report how the automation tool was used, how the tool was trained, and details on the tool's performance and internal validation. |
| EFFECT MEASURES | 12 | • Specify for each outcome (or type of outcome [e.g. binary, continuous]), the effect measure(s) (e.g. risk ratio, mean difference) used in the synthesis or presentation of results.<br>• State any thresholds (or ranges) used to interpret the size of effect (e.g. minimally important difference; ranges for no/trivial, small, moderate and large effects) and the rationale for these thresholds.<br>• If synthesized results were re-expressed to a different effect measure, report the method used to re-express results (e.g. meta-analysing risk ratios and computing an absolute risk reduction based on an assumed comparator risk).<br>• *Consider providing justification for the choice of effect measure.* |
| SYNTHESIS METHODS (eligibility for synthesis) | 13a | • Describe the processes used to decide which studies were eligible for each synthesis. |
| SYNTHESIS METHODS (preparing for synthesis) | 13b | • Report any methods required to prepare the data collected from studies for presentation or synthesis, such as handling of missing summary statistics, or data conversions. |
| SYNTHESIS METHODS (tabulation and graphical methods) | 13c | • Report chosen tabular structure(s) used to display results of individual studies and syntheses, along with details of the data presented.<br>• Report chosen graphical methods used to visually display results of individual studies and syntheses.<br>• *If studies are ordered or grouped within tables or graphs based on study characteristics (e.g. by size of the study effect, year of publication), consider reporting the basis for the chosen ordering/grouping.*<br>• *If non-standard graphs were used, consider reporting the rationale for selecting the chosen graph.* |
| SYNTHESIS METHODS (statistical synthesis methods) | 13d | • If statistical synthesis methods were used, reference the software, packages and version numbers used to implement synthesis methods.<br>• If it was not possible to conduct a meta-analysis, describe and justify the synthesis methods or summary approach used.<br>• If meta-analysis was done, specify:<br> o the meta-analysis model (fixed-effect, fixed-effects or random-effects) and provide rationale for the selected model.<br> o the method used (e.g. Mantel-Haenszel, inverse-variance).<br> o any methods used to identify or quantify statistical heterogeneity (e.g. visual inspection of results, a formal statistical test for heterogeneity, heterogeneity variance ($\tau^2$), inconsistency (e.g. $I^2$), and prediction intervals).<br>• If a random-effects meta-analysis model was used:<br> o specify the between-study (heterogeneity) variance estimator used (e.g. DerSimonian and Laird, restricted maximum likelihood (REML)).<br> o specify the method used to calculate the confidence interval for the summary effect (e.g. Wald-type confidence interval, Hartung-Knapp-Sidik-Jonkman).<br> o *consider specifying other details about the methods used, such as the method for calculating confidence limits for the heterogeneity variance.* |

| Section and Topic | Item # | Elements recommended for reporting |
|---|---|---|
| | | • If a Bayesian approach to meta-analysis was used, describe the prior distributions about quantities of interest (e.g. intervention effect being analysed, amount of heterogeneity in results across studies).<br>• If multiple effect estimates from a study were included in a meta-analysis, describe the method(s) used to model or account for the statistical dependency (e.g. multivariate meta-analysis, multilevel models or robust variance estimation).<br>• If a planned synthesis was not considered possible or appropriate, report this and the reason for that decision. |
| SYNTHESIS METHODS (methods to explore heterogeneity) | 13e | • If methods were used to explore possible causes of statistical heterogeneity, specify the method used (e.g. subgroup analysis, meta-regression).<br>• If subgroup analysis or meta-regression was performed, specify for each:<br>o which factors were explored, levels of those factors, and which direction of effect modification was expected and why (where possible).<br>o whether analyses were conducted using study-level variables (i.e. where each study is included in one subgroup only), within-study contrasts (i.e. where data on subsets of participants within a study are available, allowing the study to be included in more than one subgroup), or some combination of the above.<br>o how subgroup effects were compared (e.g. statistical test for interaction for subgroup analyses).<br>• If other methods were used to explore heterogeneity because data were not amenable to meta-analysis of effect estimates (e.g. structuring tables to examine variation in results across studies based on subpopulation), describe the methods used, along with the factors and levels.<br>• If any analyses used to explore heterogeneity were not pre-specified, identify them as such. |
| SYNTHESIS METHODS (sensitivity analyses) | 13f | • If sensitivity analyses were performed, provide details of each analysis (e.g. removal of studies at high risk of bias, use of an alternative meta-analysis model).<br>• If any sensitivity analyses were not pre-specified, identify them as such. |
| REPORTING BIAS ASSESSMENT | 14 | • Specify the methods (tool, graphical, statistical or other) used to assess the risk of bias due to missing results in a synthesis (arising from reporting biases).<br>• If risk of bias due to missing results was assessed using an existing tool, specify the methodological components/domains/items of the tool, and the process used to reach a judgement of overall risk of bias.<br>• If any adaptations to an existing tool to assess risk of bias due to missing results were made, specify the adaptations.<br>• If a new tool to assess risk of bias due to missing results was developed for use in the review, describe the content of the tool and make it publicly accessible.<br>• Report how many reviewers assessed risk of bias due to missing results in a synthesis, whether multiple reviewers worked independently, and any processes used to resolve disagreements between assessors.<br>• Report any processes used to obtain or confirm relevant information from study investigators.<br>• If an automation tool was used to assess risk of bias due to missing results, report how the automation tool was used, how the tool was trained, and details on the tool's performance and internal validation. |
| CERTAINTY ASSESSMENT | 15 | • Specify the tool or system (and version) used to assess certainty (or confidence) in the body of evidence.<br>• Report the factors considered (e.g. precision of the effect estimate, consistency of findings across studies) and the criteria used to assess each factor when assessing certainty in the body of evidence.<br>• Describe the decision rules used to arrive at an overall judgement of the level of certainty, together with the intended interpretation (or definition) of each level of certainty.<br>• If applicable, report any review-specific considerations for assessing certainty, such as thresholds used to assess imprecision and ranges of magnitude of effect that might be considered trivial, moderate or large, and the rationale for these thresholds and ranges (item #12). |

| Section and Topic | Item # | Elements recommended for reporting |
|---|---|---|
| | | • If any adaptations to an existing tool or system to assess certainty were made, specify the adaptations.<br>• Report how many reviewers assessed certainty in the body of evidence for an outcome, whether multiple reviewers worked independently, and any processes used to resolve disagreements between assessors.<br>• Report any processes used to obtain or confirm relevant information from investigators.<br>• If an automation tool was used to support the assessment of certainty, report how the automation tool was used, how the tool was trained, and details on the tool's performance and internal validation.<br>• Describe methods for reporting the results of assessments of certainty, such as the use of Summary of Findings tables.<br>• If standard phrases that incorporate the certainty of evidence were used (e.g. "hip protectors *probably* reduce the risk of hip fracture slightly"), report the intended interpretation of each phrase and the reference for the source guidance. |
| **RESULTS** | | |
| STUDY SELECTION (flow of studies) | 16a | • Report, ideally using a flow diagram, the number of: records identified; records excluded before screening; records screened; records excluded after screening titles or titles and abstracts; reports retrieved for detailed evaluation; potentially eligible reports that were not retrievable; retrieved reports that did not meet inclusion criteria and the primary reasons for exclusion; and the number of studies and reports included in the review. If applicable, also report the number of ongoing studies and associated reports identified.<br>• If the review is an update of a previous review, report results of the search and selection process for the current review and specify the number of studies included in the previous review.<br>• If applicable, indicate in the PRISMA flow diagram how many records were excluded by a human and how many by automation tools. |
| STUDY SELECTION (excluded studies) | 16b | • Cite studies that might appear to meet the inclusion criteria, but which were excluded, and explain why they were excluded. |
| STUDY CHARACTERISTICS | 17 | • Cite each included study.<br>• Present the key characteristics of each study in a table or figure (considering a format that will facilitate comparison of characteristics across the studies).<br>• *If the review examines the effects of interventions, consider presenting an additional table that summarises the intervention details for each study.* |
| RISK OF BIAS IN STUDIES | 18 | • Present tables or figures indicating for each study the risk of bias in each domain/component/item assessed (e.g. blinding of outcome assessors, missing outcome data) and overall study-level risk of bias.<br>• Present justification for each risk of bias judgement, for example in the form of relevant quotations from reports of included studies.<br>• *If assessments of risk of bias were done for specific outcomes or results in each study, consider displaying risk of bias judgements on a forest plot, next to the study results.* |
| RESULTS OF INDIVIDUAL STUDIES | 19 | • For all outcomes, irrespective of whether statistical synthesis was undertaken, present for each study summary statistics for each group (where appropriate). For dichotomous outcomes, report the number of participants with and without the events for each group; or the number with the event and the total for each group (e.g. 12/45). For continuous outcomes, report the mean, standard deviation and sample size of each group.<br>• For all outcomes, irrespective of whether statistical synthesis was undertaken, present for each study an effect estimate and its precision (e.g. standard error or 95% confidence/credible interval). For example, for time-to-event outcomes, present a hazard ratio and its confidence interval.<br>• If study-level data is presented visually or reported in the text (or both), also present a tabular display of the results.<br>• If results were obtained from multiple data sources (e.g. journal article, study register entry, clinical study report, correspondence with authors), report the source of the data.<br>• If applicable, indicate which results were not reported directly and had to be computed or estimated from other information. |

| Section and Topic | Item # | Elements recommended for reporting |
|---|---|---|
| RESULTS OF SYNTHESES (characteristics of contributing studies) | 20a | • Provide a brief summary of the characteristics and risk of bias among studies contributing to each synthesis (meta-analysis or other). The summary should focus only on study characteristics that help in interpreting the results (especially those that suggest the evidence addresses only a restricted part of the review question, or indirectly addresses the question).<br>• Indicate which studies were included in each synthesis (e.g. by listing each study in a forest plot or table or citing studies in the text). |
| RESULTS OF SYNTHESES (results of statistical syntheses) | 20b | • Report results of all statistical syntheses described in the protocol and all syntheses conducted that were not pre-specified.<br>• If meta-analysis was conducted, report for each:<br>o the summary estimate and its precision (e.g. standard error or 95% confidence/credible interval)<br>o measures of statistical heterogeneity (e.g. $\tau^2$, $I^2$, prediction interval)<br>• If other statistical synthesis methods were used (e.g. summarising effect estimates, combining P values), report the synthesized result and a measure of precision (or equivalent information, for example, the number of studies and total sample size).<br>• If the statistical synthesis method does not yield an estimate of effect (e.g. as is the case when P values are combined), report the relevant statistics (e.g. P value from the statistical test), along with an interpretation of the result that is consistent with the question addressed by the synthesis method.<br>• If comparing groups, describe the direction of effect (e.g. fewer events in the intervention group, or higher pain in the comparator group).<br>• If synthesising mean differences, specify for each synthesis, where applicable, the unit of measurement (e.g. kilograms or pounds for weight), the upper and lower limits of the measurement scale (e.g. anchors range from 0 to 10), direction of benefit (e.g. higher scores denote higher severity of pain), and the minimally important difference, if known. If synthesising standardised mean differences, and the effect estimate is being re-expressed to a particular instrument, details of the instrument, as per the mean difference, should be reported. |
| RESULTS OF SYNTHESES (results of investigations of heterogeneity) | 20c | • If investigations of possible causes of heterogeneity were conducted:<br>o present results regardless of the statistical significance, magnitude, or direction of effect modification.<br>o identify the studies contributing to each subgroup.<br>o report results with due consideration to the observational nature of the analysis and risk of confounding due to other factors.<br>• If subgroup analysis was conducted:<br>o report for each analysis the exact P value for a test for interaction, as well as, within each subgroup, the summary estimates, their precision (e.g. standard error or 95% confidence/credible interval) and measures of heterogeneity.<br>o *consider presenting the estimate for the difference between subgroups and its precision.*<br>• If meta-regression was conducted:<br>o report for each analysis the exact P value for the regression coefficient and its precision.<br>o *consider presenting a meta-regression scatterplot with the study effect estimates plotted against the potential effect modifier.*<br>• If informal methods (i.e. those that do not involve a formal statistical test) were used to investigate heterogeneity, describe the results observed. |
| RESULTS OF SYNTHESES (results of sensitivity analyses) | 20d | • If any sensitivity analyses were conducted:<br>o report the results for each sensitivity analysis.<br>o comment on how robust the main analysis was given the results of all corresponding sensitivity analyses.<br>o *consider presenting results in tables that indicate: (i) the summary effect estimate, a measure of precision (and potentially other relevant statistics, for example, $I^2$ statistic) and contributing studies for the original meta-analysis; (ii) the same information for the sensitivity analysis; and (iii) details of the original and sensitivity analysis assumptions.*<br>o *consider presenting results of sensitivity analyses visually using forest plots.* |

*From:* Page MJ, McKenzie JE, Bossuyt PM, Boutron I, Hoffmann TC, Mulrow CD, et al. The PRISMA 2020 statement: an updated guideline for reporting systematic reviews. BMJ 2021;372:n71. doi: 10.1136/bmj.n71
For more information, visit: http://www.prisma-statement.org/

| Section and Topic | Item # | Elements recommended for reporting |
|---|---|---|
| REPORTING BIASES | 21 | • Present assessments of risk of bias due to missing results (arising from reporting biases) for each synthesis assessed.<br>• If a tool was used to assess risk of bias due to missing results in a synthesis, present responses to questions in the tool, judgements about risk of bias and any information used to support such judgements.<br>• If a funnel plot was generated to evaluate small-study effects (one cause of which is reporting biases), present the plot and specify the effect estimate and measure of precision used in the plot. If a contour-enhanced funnel plot was generated, specify the 'milestones' of statistical significance that the plotted contour lines represent (P = 0.01, 0.05, 0.1, etc.)<br>• If a test for funnel plot asymmetry was used, report the exact P value observed for the test, and potentially other relevant statistics, for example the standardised normal deviate, from which the P value is derived.<br>• If any sensitivity analyses seeking to explore the potential impact of missing results on the synthesis were conducted, present results of each analysis (see item #20d), compare them with results of the primary analysis, and report results with due consideration of the limitations of the statistical method.<br>• *If studies were assessed for selective non-reporting of results by comparing outcomes and analyses pre-specified in study registers, protocols, and statistical analysis plans with results that were available in study reports, consider presenting a matrix (with rows as studies and columns as syntheses) to present the availability of study results.*<br>• *If an assessment of selective non-reporting of results reveals that some studies are missing from the synthesis, consider displaying the studies with missing results underneath a forest plot or including a table with the available study results.* |
| CERTAINTY OF EVIDENCE | 22 | • Report the overall level of certainty (or confidence) in the body of evidence for each important outcome.<br>• Provide an explanation of reasons for rating down (or rating up) the certainty of evidence (e.g. in footnotes to an evidence summary table).<br>• Communicate certainty in the evidence wherever results are reported (i.e. abstract, evidence summary tables, results, conclusions), using a format appropriate for the section of the review.<br>• *Consider including evidence summary tables, such as GRADE Summary of Findings tables.* |
| **DISCUSSION** | | |
| DISCUSSION (interpretation) | 23a | • Provide a general interpretation of the results in the context of other evidence. |
| DISCUSSION (limitations of evidence) | 23b | • Discuss any limitations of the evidence included in the review. |
| DISCUSSION (limitations of review processes) | 23c | • Discuss any limitations of the review processes used, and comment on the potential impact of each limitation. |
| DISCUSSION (implications) | 23d | • Discuss implications of the results for practice and policy.<br>• Make explicit recommendations for future research. |
| **OTHER INFORMATION** | | |
| REGISTRATION AND PROTOCOL (registration) | 24a | • Provide registration information for the review, including register name and registration number, or state that the review was not registered. |
| REGISTRATION AND PROTOCOL (protocol) | 24b | • Indicate where the review protocol can be accessed (e.g. by providing a citation, DOI or link), or state that a protocol was not prepared. |

*From:* Page MJ, McKenzie JE, Bossuyt PM, Boutron I, Hoffmann TC, Mulrow CD, et al. The PRISMA 2020 statement: an updated guideline for reporting systematic reviews. BMJ 2021;372:n71. doi: 10.1136/bmj.n71
For more information, visit: http://www.prisma-statement.org/

| Section and Topic | Item # | Elements recommended for reporting |
|---|---|---|
| REGISTRATION AND PROTOCOL (amendments) | 24c | • Report details of any amendments to information provided at registration or in the protocol, noting: (a) the amendment itself; (b) the reason for the amendment; and (c) the stage of the review process at which the amendment was implemented. |
| SUPPORT | 25 | • Describe sources of financial or non-financial support for the review, specifying relevant grant ID numbers for each funder. If no specific financial or non-financial support was received, this should be stated.<br>• Describe the role of the funders or sponsors (or both) in the review. If funders or sponsors had no role in the review, this should be declared. |
| COMPETING INTERESTS | 26 | • Disclose any of the authors' relationships or activities that readers could consider pertinent or to have influenced the review.<br>• If any authors had competing interests, report how they were managed for particular review processes. |
| AVAILABILITY OF DATA, CODE, AND OTHER MATERIALS | 27 | • Report which of the following are publicly available: template data collection forms; data extracted from included studies; data used for all analyses; analytic code; any other materials used in the review.<br>• If any of the above materials are publicly available, report where they can be found (e.g. provide a link to files deposited in a public repository).<br>• If data, analytic code, or other materials will be made available upon request, provide the contact details of the author responsible for sharing the materials and describe the circumstances under which such materials will be shared. |

*From:* Page MJ, McKenzie JE, Bossuyt PM, Boutron I, Hoffmann TC, Mulrow CD, et al. The PRISMA 2020 statement: an updated guideline for reporting systematic reviews. BMJ 2021;372:n71. doi: 10.1136/bmj.n71
For more information, visit: http://www.prisma-statement.org/
